# Evaluation of commercially available high-throughput SARS-CoV-2 serological assays for serosurveillance and related applications

**DOI:** 10.1101/2021.09.04.21262414

**Authors:** Mars Stone, Eduard Grebe, Hasan Sulaeman, Clara Di Germanio, Honey Dave, Kathleen Kelly, Brad Biggerstaff, Bridgit O. Crews, Nam Tran, Keith R. Jerome, Thomas N. Denny, Boris Hogema, Mark Destree, Jefferson M. Jones, Natalie Thornburg, Graham Simmons, Mel Krajden, Steve Kleinman, Larry J. Dumont, Michael P. Busch

**Author notes:** Corresponding author: Mars Stone, PhD., Vitalant Research Institute, San Francisco CA, USA, Department of Laboratory Medicine, University of California San Francisco, CA, USA. 270 Masonic Ave, San Francisco, CA 94118. These first authors contributed equally to this article (Mars Stone, Eduard Grebe). Disclaimer: The findings and conclusions in this report are those of the authors and do not necessarily represent the official position of the Centers for Disease Control and Prevention.

## Abstract

SARS-CoV-2 serosurveys can estimate cumulative incidence for monitoring epidemics but require characterization of employed serological assays performance to inform testing algorithm development and interpretation of results. We conducted a multi-laboratory evaluation of 21 commercial high-throughput SARS-CoV-2 serological assays using blinded panels of 1,000 highly-characterized blood-donor specimens. Assays demonstrated a range of sensitivities (96%-63%), specificities (99%-96%) and precision (IIC 0.55-0.99). Durability of antibody detection in longitudinal samples was dependent on assay format and immunoglobulin target, with anti-spike, direct, or total Ig assays demonstrating more stable, or increasing reactivity over time than anti-nucleocapsid, indirect, or IgG assays. Assays with high sensitivity, specificity and durable antibody detection are ideal for serosurveillance. Less sensitive assays demonstrating waning reactivity are appropriate for other applications, including characterizing antibody responses after infection and vaccination, and detection of anamnestic boosting by reinfections and vaccine breakthrough infections. Assay performance must be evaluated in the context of the intended use.

## Introduction

Serosurveillance for SARS-CoV-2 infection is critical to monitor the course of the evolving pandemic and local outbreaks, and informs infection fatality ratios, vaccine penetrance and the impact of mitigation measures, and levels of population immunity. Serosurveillance should be conducted with representative population sampling using well characterized serological assays selected based on their performance characteristics and optimized algorithms. The use of assays and algorithms that detect mild or asymptomatic infections are critical for accurately estimating cumulative incidence, and case- and death-to-infection ratios.

More than >85 SARS-CoV-2 antibody (Ab) assays received FDA Emergency Use Authorization (EUA) as of August 19, 2021, ranging from point-of-care tests to fully automated high-throughput platforms [1]. These assays target different immunoglobulins (Ig) against viral antigens (full length Spike protein [S1/S2], subunit 1 [S1] and/or subunit 2 [S2] of Spike, the receptor binding domain [RBD] of Spike, or the nucleocapsid protein [N]). Detection methods include lateral flow assays [LFA], enzyme-linked immunosorbent assays [ELISA], and chemiluminescent immunoassay [CLIA], and detection of either total Ig, or selective IgG, IgM or IgA antibodies [1]. There are limited head-to-head evaluation data available for high-throughput SARS-CoV-2 serological assays and few large-scale studies that have focused on performance for serosurveillance applications. Comprehensive characterization of assay performance must include sensitivity, specificity, and durability of antibody detection over time since infection.

We conducted a standardized, multi-laboratory comparative assessment of 21 high-throughput, commercially available SARS-CoV-2 serological assays using blinded panels of 1,000 highly characterized de-identified specimens including longitudinal and cross sectional COVID-19 convalescent plasma (CCP) and pre-COVID control plasma specimens. Panels were distributed to experienced testing laboratories determined to be proficient by the manufacturers. Assays were selected to represent multiple formats and antigen targets. Data from this study will inform assay selection and development of testing algorithms to meet the optimal performance characteristics for primary screening and supplemental testing in US and global serosurveillance studies. The study also provides performance data relevant to other serological testing contexts that will allow clinicians, public health organizations, laboratorians, and emergency response planners to develop optimal algorithms for infection detection and confirmation, including vaccine breakthrough and recurrent infections.

## Methods

### Assay selection, panel development and testing

Key characteristics of the assays included, such as format and configuration, antigen composition, and immunoglobulin target, are summarized in Table 1. Uniquely blinded identical panels were distributed to experienced testing laboratories to determine performance characteristics including sensitivity, specificity, repeatability, dilutional performance, and durability of reactivity over time.

**Table 1.**
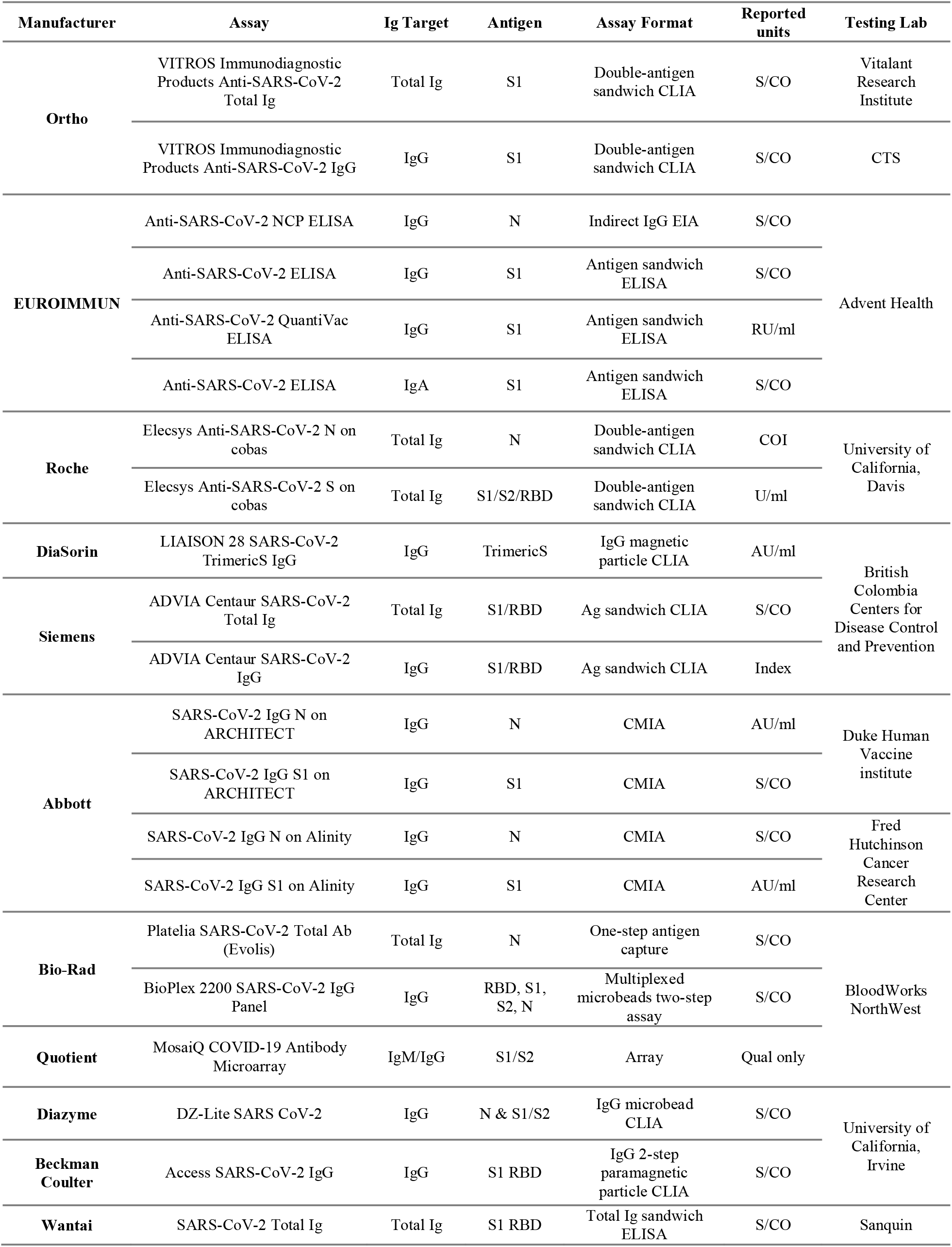
Serological assays and key characteristics. S, spike protein; RBD, receptor binding domain; N, nucleocapsid; Ag, antigen; Ab, antibody; Ig, immunoglobulin; S/CO, signal to cutoff ratio; EIA, enzyme immunoassay; ELISA, enzyme-linked immunosorbent assay;RU, relative units, AU, arbitrary units; CLIA, Chemiluminescent immunoassay; CMIA, Chemiluminescent microparticle immunoassay. Current US regulatory status can be obtained on the FDA website at: https://www.fda.gov/medical-devices/coronavirus-disease-2019-covid-19-emergency-use-authorizations-medical-devices/in-vitro-diagnostics-euas-serology-and-other-adaptive-immune-response-tests-sars-cov-2.

Plasma or serum specimens were obtained from apheresis plasma units or associated whole blood tubes from consenting CCP donors from March through November 2020. Specimens were shipped from collection sites and stored frozen at Vitalant Research Institute (VRI) until panel assembly and frozen distribution. All blood donors consented to use of de-identified, residual specimens for further research purposes. Consistent with the policies and guidance of the UCSF IRB, VRI self-certified that use of the de-identified CCP donations in this study does not meet the criteria for human subjects research. CDC investigators reviewed and relied on this determination as consistent with applicable federal law and CDC policy (45 C.F.R. part 46, 21 C.F.R. part 56; 42 U.S.C. Sect. 241(d); 5 U.S.C. Sect. 552a; 44 U.S.C. Sect. 3501). Qualification for CCP donation required documentation of positive SARS-CoV-2 molecular or serologic test, complete resolution of symptoms 14-28 days prior to donation[2], and reactivity on the primary screening Ortho VITROS SARS-CoV-2 S total Ig (Ortho Clinical Diagnostics, Raritan, NJ) Ab assay and standard allogeneic blood donor qualification criteria[3].

To evaluate the waning of sensitivity over time, longitudinal specimens were included from 24 CCP donors who continued to qualify for CCP donation at each of 4-14 donations (median 9) over 79-126 days (median 95). A COVID-19 Seroconversion Panel consisted of 14 time points from a single source plasma donor during the progression of a SARS-CoV-2 infection over 87 days[4]. Fifteen CCP specimens were represented in 6 blinded replicates to evaluate precision. The dilution panel consisted of six 4-fold serial dilutions of specimens with a range of neat Ab titers [5]. The panel also included 24 apparent serosilent specimens from donors who initially qualified for CCP donation as having a positive molecular test but without evidence of seroconversion by the Ortho S total Ig assay. Specificity panel included pre-pandemic blood donor specimens derived from plasma components collected before end of year 2019 (N=432) and 27 donations collected in early 2020 that tested non-reactive on Ortho CoV2T and were non-neutralizing by pseudovirus neutralization assay [5].

### Statistical Analysis

All statistical analyses were performed using the R statistical programming language (v. 4.0.4, [6]) and using various packages, including the *binom* package for confidence intervals on proportions [7], the *glm2* package [8] for regression analysis and the *ggplot2* package [9] for plotting.

### Sensitivity and Specificity

Sensitivity was assessed in cross sectional CCP specimens. Because data on symptoms, clinical severity, hospitalization, and diagnostic test results (molecular or antigen) were not available, we defined “true positivity” according to three sets of criteria: (1) qualification as a CCP donor according to blood center policies, which required donors to provide evidence of a SARS-CoV-2 diagnosis, with symptomatic infection resolved at least 2 weeks prior to the first donation (N=191), [https://www.fda.gov/media/141477/download], (2) confirmation of detectable nAb by the Broad live virus neutralization assay [5] (N=154), or (3) reactive on ≥3 evaluated binding Ab (bAb) tests (N=198). There is substantial overlap between the three definitions, with 149 specimens classified as positive by all three definitions, 34 by (1) and (3), 3 by (2) and (3) and only 22 positive by only one of the definitions. The 24 purposely selected “serosilent” CCP specimens (see Table 2) were excluded from the sensitivity analysis based on criterion 1 above, while specimens from the longitudinal CCP donor cohort were excluded from all sensitivity analyses. Donors who continued to qualify for CCP donation may bias sensitivity estimates given they were required to have bAb reactivity for continued donation of CCP.

**Table 2.**
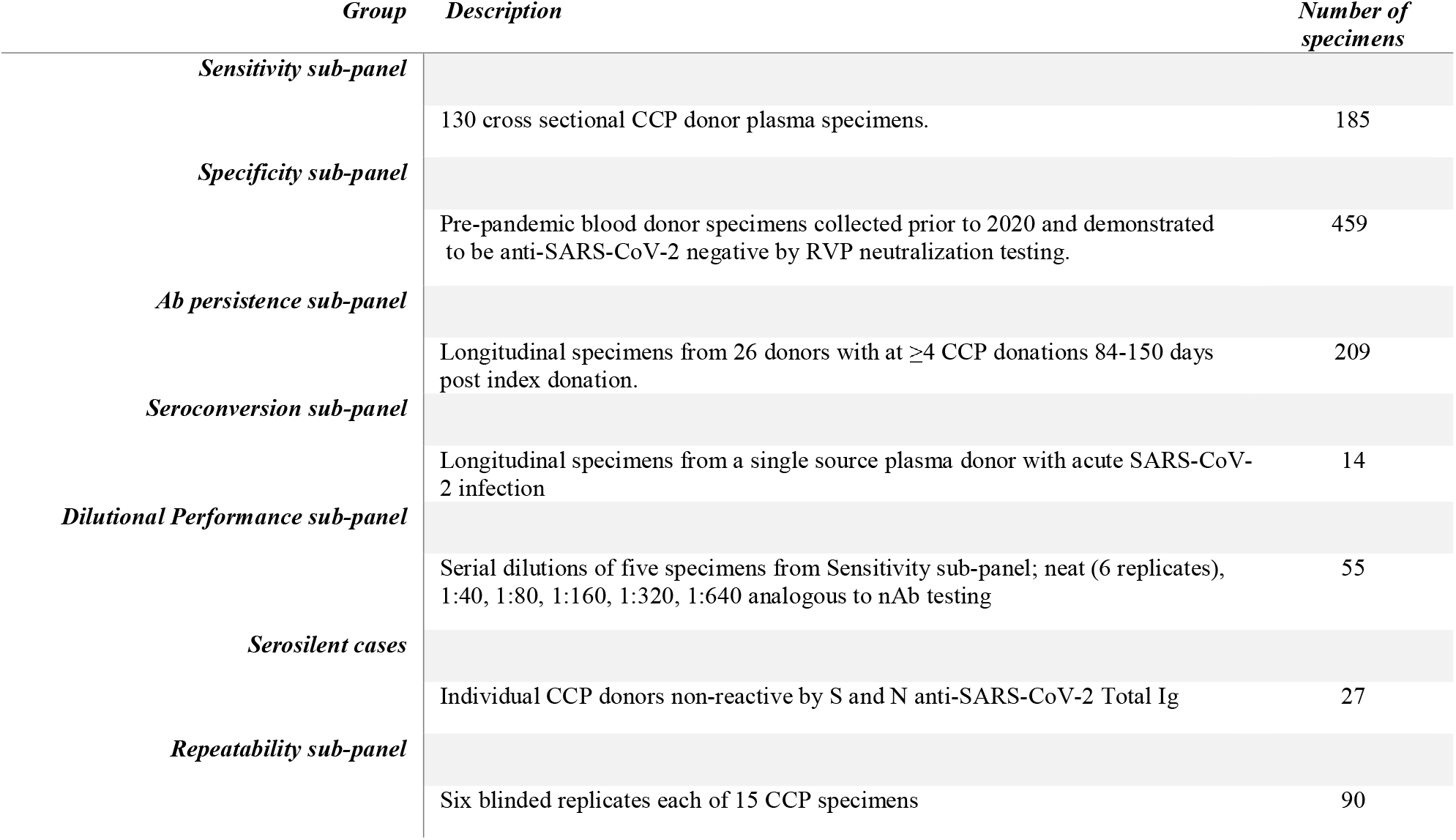
Composition of the assessment panel. CCP, COVID convalescent plasma, S, spike protein; RBD, receptor binding domain; N, nucleocapsid; Ag, antigen; Ab, antibody; Ig, immunoglobulin; RVP, reporter viral particle; bAb, neutralizing antibodies.

Specificity was assessed using pre-pandemic blood donor specimens (N=432) and 27 seronegative early 2020 donations [5]. These 27 samples were not included in the primary analysis but were included in a secondary specificity analysis (N=459) (Appendix Figure 1).

All sensitivity and specificity estimates were based on reported qualitative interpretations of assay results. Results defined and reported by the manufacturer as “equivocal” were excluded from primary sensitivity and specificity estimates. A secondary analysis of sensitivity was conducted in which we considered results reported as equivocal by the testing lab as non-reactive (Appendix Figure 2). All 95% confidence intervals are Wilson score intervals.

### Repeatability and Assay Precision

Coefficients of variation (CV, i.e., the ratio of the standard deviation across measurements of the six replicate specimens to the mean of the six measurements, expressed as a percentage) were computed for each of the replicate specimens (N=90). A limitation of this approach is that assays with narrower dynamic range produced very low or zero CVs for results at the upper limit of quantification. To adequately account for the impact of specimens with reactivity outside the measurement range, we excluded these specimens from the overall repeatability assessment for which we used the intraclass correlation coefficients (ICCs). The ICC expresses between-sample variance as a proportion of total variance in the tested replicate specimen. In the case of the Bio-Rad BioPlex assay (Bio-Rad, Hercules, CA), on-board dilutions were conducted by the testing lab and used to estimate reactivity in specimens where initial results were above the assay’s limit of quantitation.

### Dilutional performance

The dilution panel (N=55) allows comparative assessment of the linearity of observed vs. expected reactivity measurements above and below assay cutoffs. Expected reactivity is defined as the mean signal intensity measured over six replicates of the neat specimen divided by the dilution factor. These analyses are reported in supplemental materials.

### Durability of antibody detection

We assessed both qualitative and quantitative durability of bAb detection in longitudinal CCP specimens (N=209 specimens from 27 donors). Documented dates of symptom onset, symptom resolution or nucleic acid test (NAT)-based diagnosis are not available for these donors, so all analyses are anchored to the index donation. These CCP donors first presented for donation early in the pandemic, typically within one month of symptom resolution [5].

Qualitative detection was assessed by estimating the proportion of specimens with detectable bAbs grouped in 30-day bins of time since index donations. To account for within-donor correlation, if a donor contributed more than one specimen in a particular time bin, the proportion of the donor’s specimens that were reactive was added to the numerator for the bin, and only 1 to the denominator, so that the proportion detected reported is the proportion of donors whose bAbs could be detected in each bin.

Quantitative detection was assessed by fitting linear mixed effects regression models with time since index donation as the predictor. Average (fixed) slopes from models fit to log-transformed and rescaled signal intensities to a fixed range, and assay signal half-lives were estimated. (Appendix 1).

## Results

When a true positive was defined by qualification as a CCP donor, the lowest assay sensitivity was 63.6% (95% CI: 56.3%-70.4%; EUROIMMUN IgA assay), and the highest was 95.8% (95% CI: 92.0%-7.9%; Ortho VITROS Total Ig S assay; Figure 1, panel A). When a true positive was defined by confirmed neutralization by plaque reduction neutralization test (PRNT), the lowest assay sensitivity was 69.7% (95% CI: 61.7%-76.7%; EUROIMMUN IgA assay), and the highest was 98.7% (95% CI: 95.4%-99.6%; Ortho VITROS Total Ig S assay; Figure 1, panel B). Most assays (17/20) had sensitivities >80%, 12/20 had sensitivities >90%, and 7/20 had sensitivities >95%, by the first definition. No assays reached a sensitivity of 96% by CCP qualification criteria or 99% by detectable nAbs criteria. Assays with the lowest sensitivity were the Beckman Coulter Access SARS-CoV-2 IgG, Diazyme DZ-Lite SARS-CoV-2 IgG and EUROIMMUN IgA assays, with estimates below 80%. Figure 1, panel C shows similar patterns to the first and second definitions of true positivity when true positivity was defined by the ‘operational standard’ of positivity based on bAb reactivity on three or more assays.

**Figure 1.**
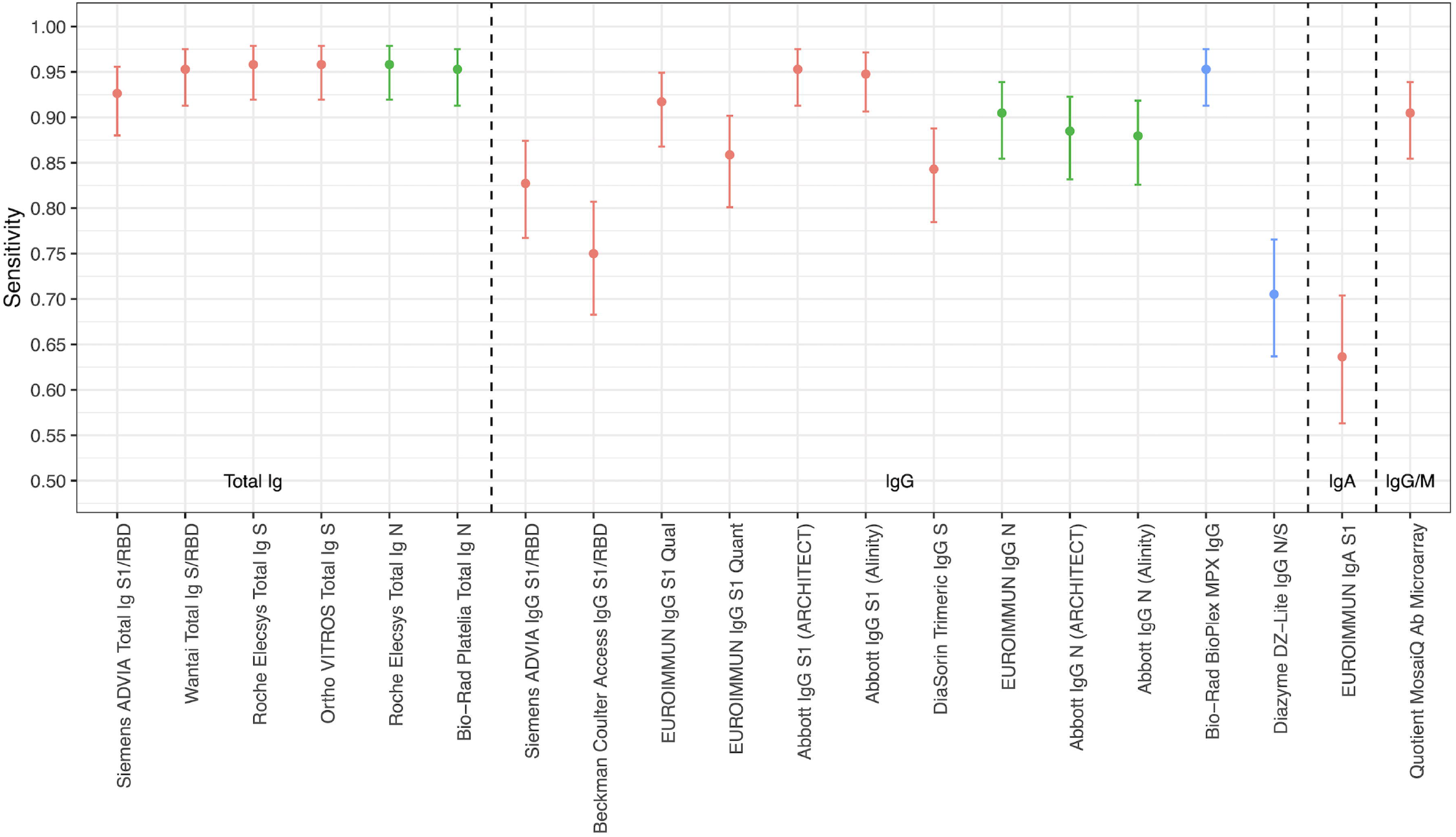
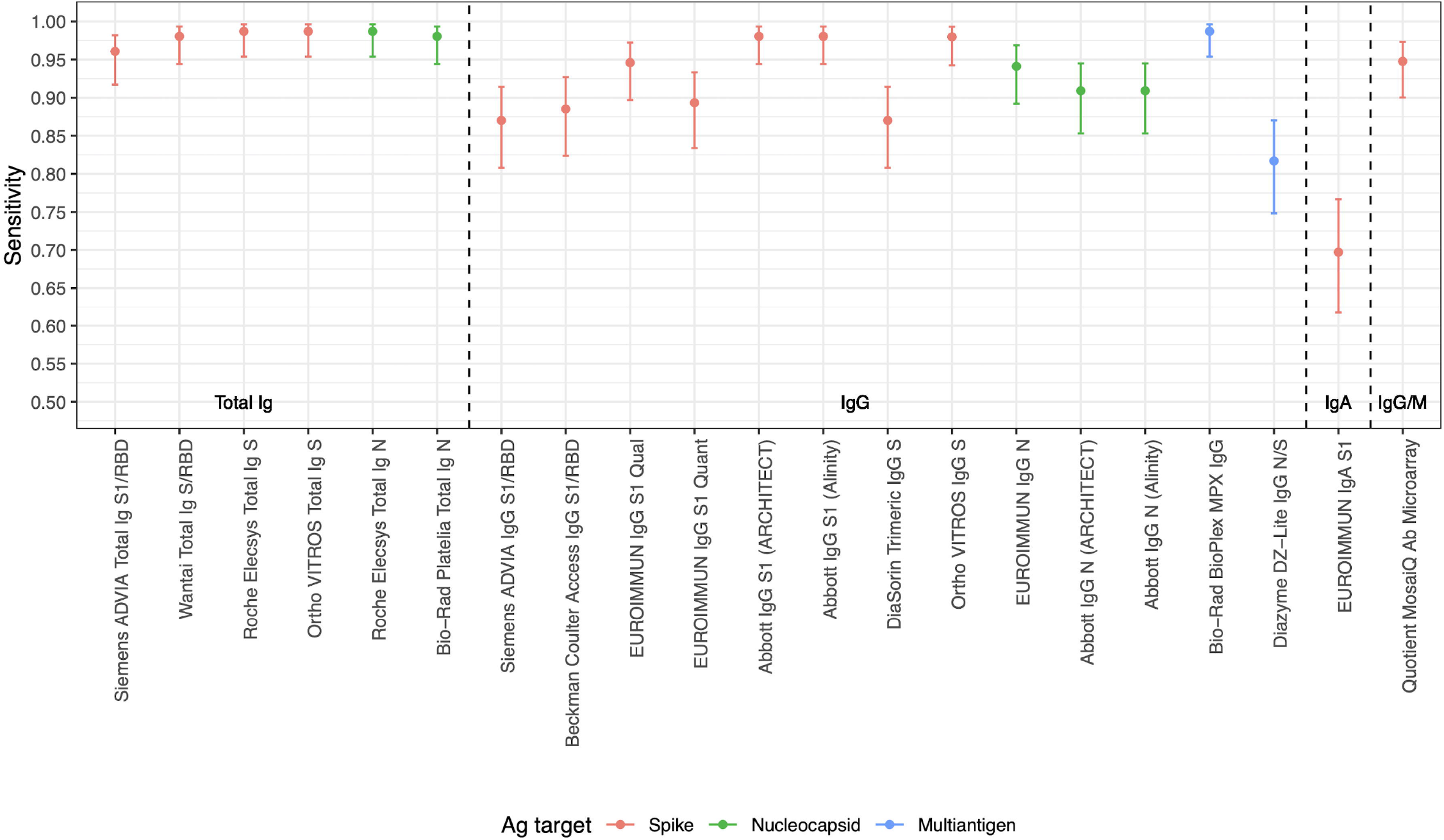
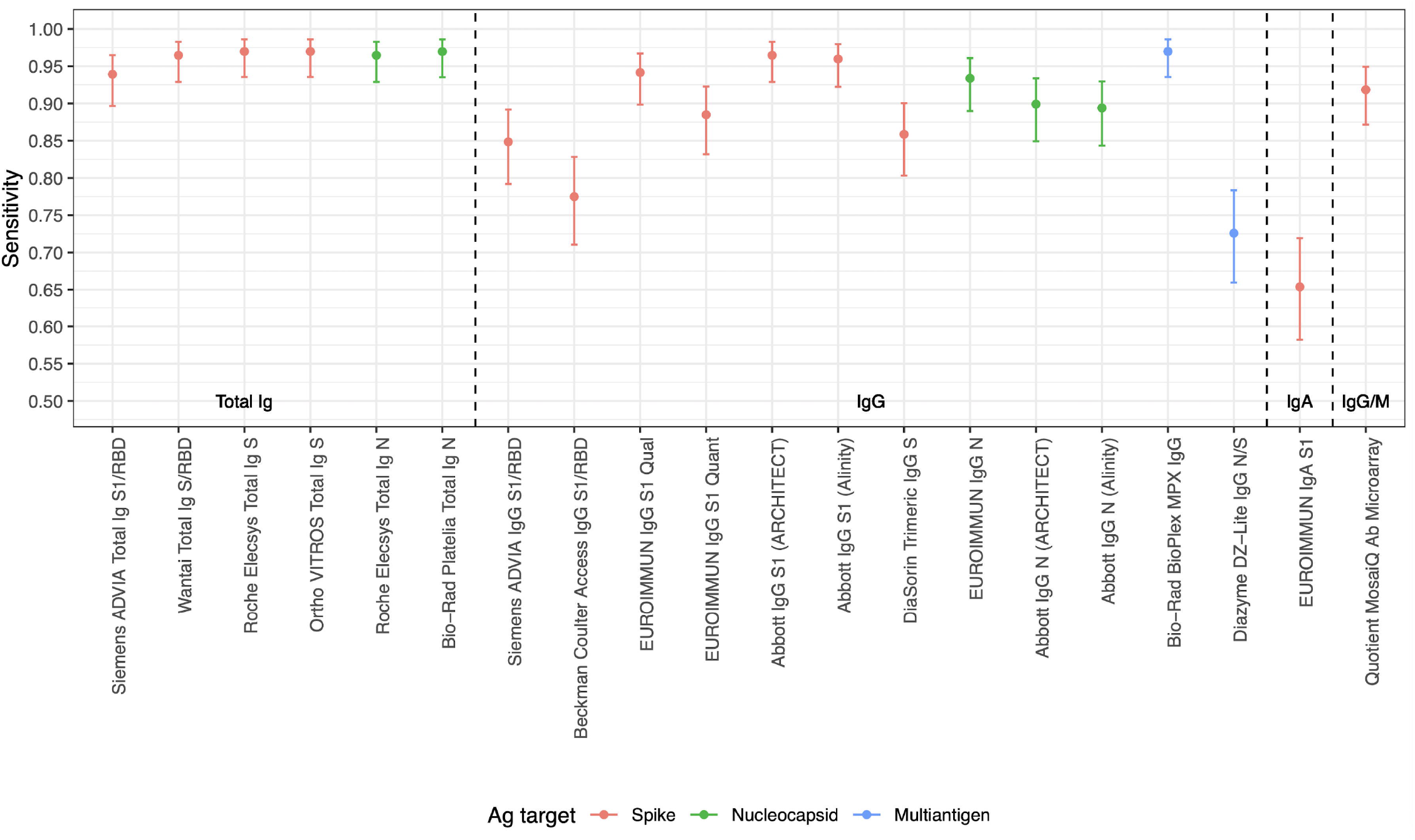
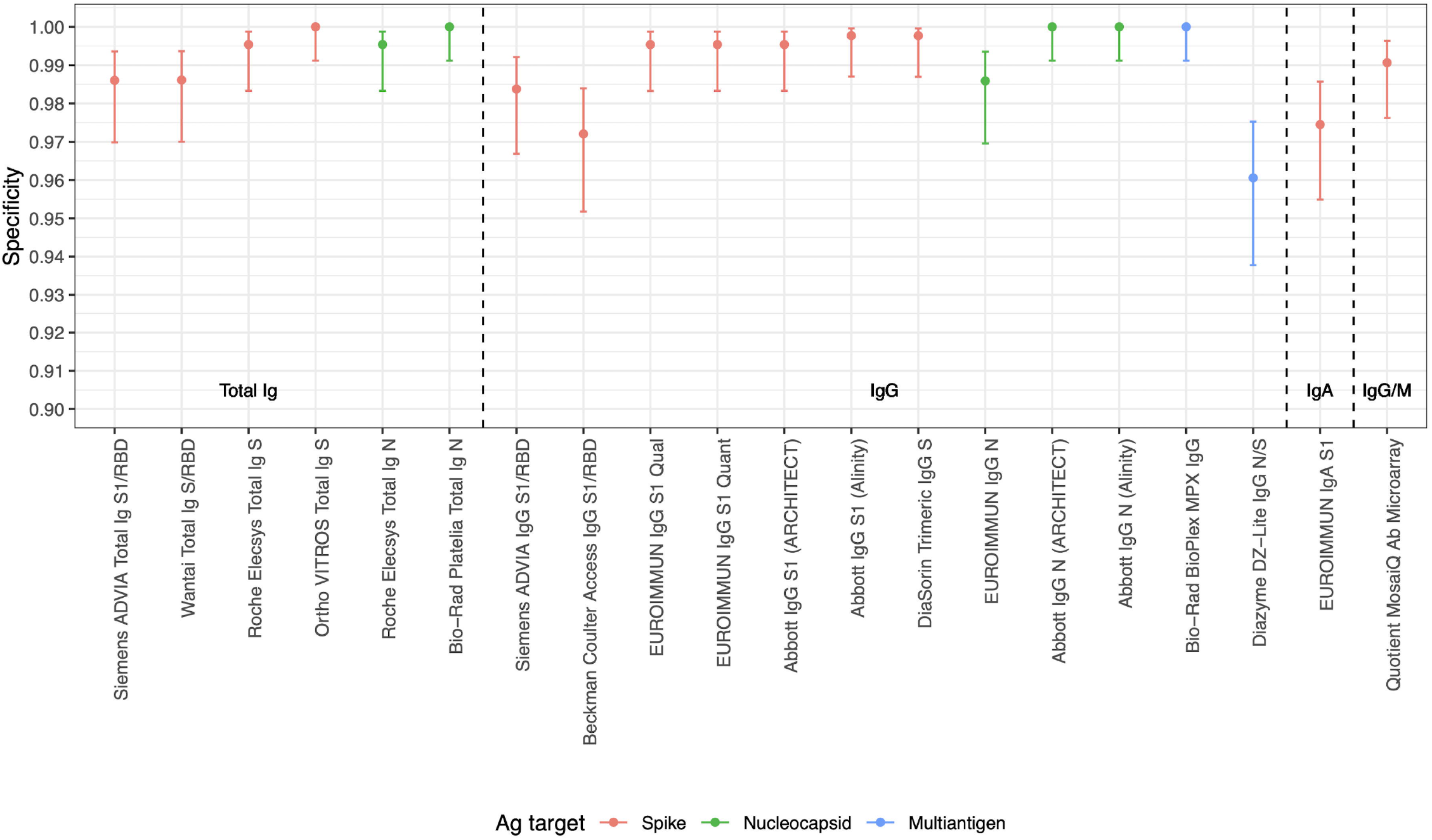
Sensitivity of SARS-CoV-2 serological assays using three definitions of a “true positive.” Panel A: Positivity defined by qualification as CCP donor (excluding purposely selected serosilents) Panel B: Positivity defined by neutralizing activity measured by Broad PRNT Panel C: Positivity defined by ‘operational standard’ (3 or more bAb assays reactive). Dots indicate point estimates and bars indicate Wilson score 95% confidence intervals. *The Ortho VITROS IgG S assay is included only in panel B, because the assay required use of serum for testing, thus only specimens with available serum and neutralizing data were tested*. S, spike protein; RBD, receptor binding domain; N, nucleocapsid; Ag, antigen; Ab, antibody; Ig, immunoglobulin; PRNT, plaque reduction neutralization test. See Table 1 for assay details.

Specificities, based on testing 432 pre-COVID-19 specimens, were high, with estimates ranging from 96.1% (95%CI: 93.8%-7.5%; Diazyme DZ-Lite assay) to 100% (95%CI: 99.1%-100%; Abbott IgG N, Bio-Rad BioPlex IgG, Bio-Rad Platelia Total Ig N, and Ortho VITROS Total Ig S assays. Most assays (13/20) had specificities above 99%, and 5/20 assays had specificities of 100% in this panel (Figure 2). Assays with poorer specificity tended to have poorer sensitivity, suggesting no tradeoff between sensitivity and specificity (Appendix Table 1 and Appendix Figure 3). Specificity estimates that included the 27 specimens from 2020 are shown in Appendix Figure 1. Secondary sensitivity analysis with ‘equivocal’ results categorized as non-reactive is shown in Appendix Figure 2. The inclusion of these 2020 and equivocal samples had minimal impact on estimates of specificity and sensitivity respectively.

**Figure 2.**
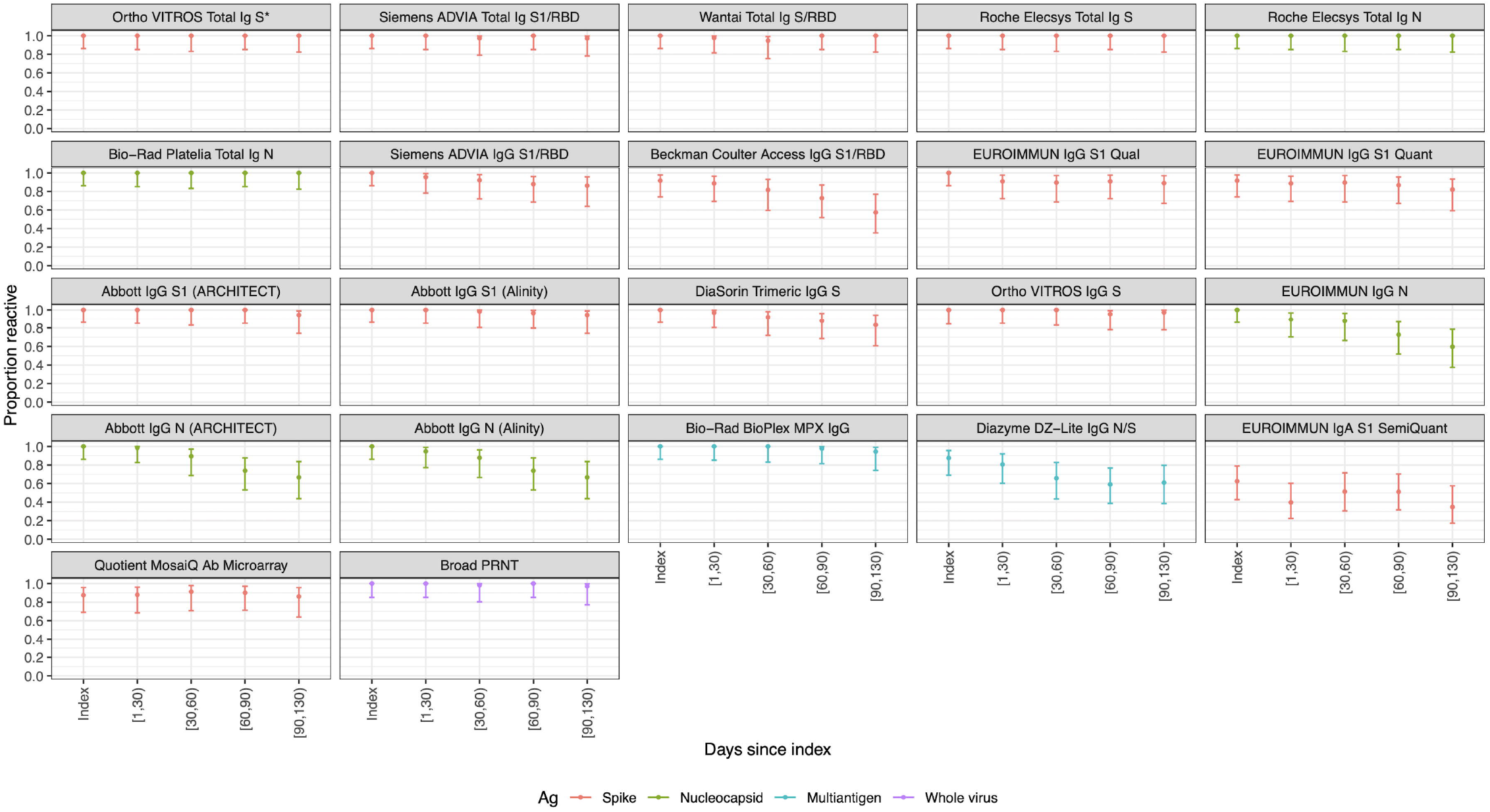
Specificity of SARS-CoV-2 serological assays in pre-COVID-19 negative control specimens.* *Dots indicate point estimates and bars indicate Wilson score 95 confidence intervals. S, spike protein; RBD, receptor binding domain; N, nucleocapsid; Ag, antigen; Ab, antibody; Ig, immunoglobulin. See Table 1 for assay details.

Durability of bAb detection was highly variable, with some assays reactive at all longitudinal timepoints, while others showed substantial declines in the proportion of reactive specimens over time (Figure 3). IgG assays and anti-N assays generally demonstrated more rapid seroreversion proportions compared to total Ig and anti-S assays. For example, the Abbott and EUROIMMUN IgG anti-N assays detected antibodies in <70% of specimens collected >90 days after index donation, while total Ig assays like the Ortho Vitros S total Ig and Roche Elecsys N total Ig assays detected antibodies in 100% of specimens at these timepoints. Given the relatively small number of donors in the cohort, the declining detection rates at later timepoints were generally not statistically distinguishable from sensitivity at earlier timepoints for these qualitative assays.

**Figure 3:**
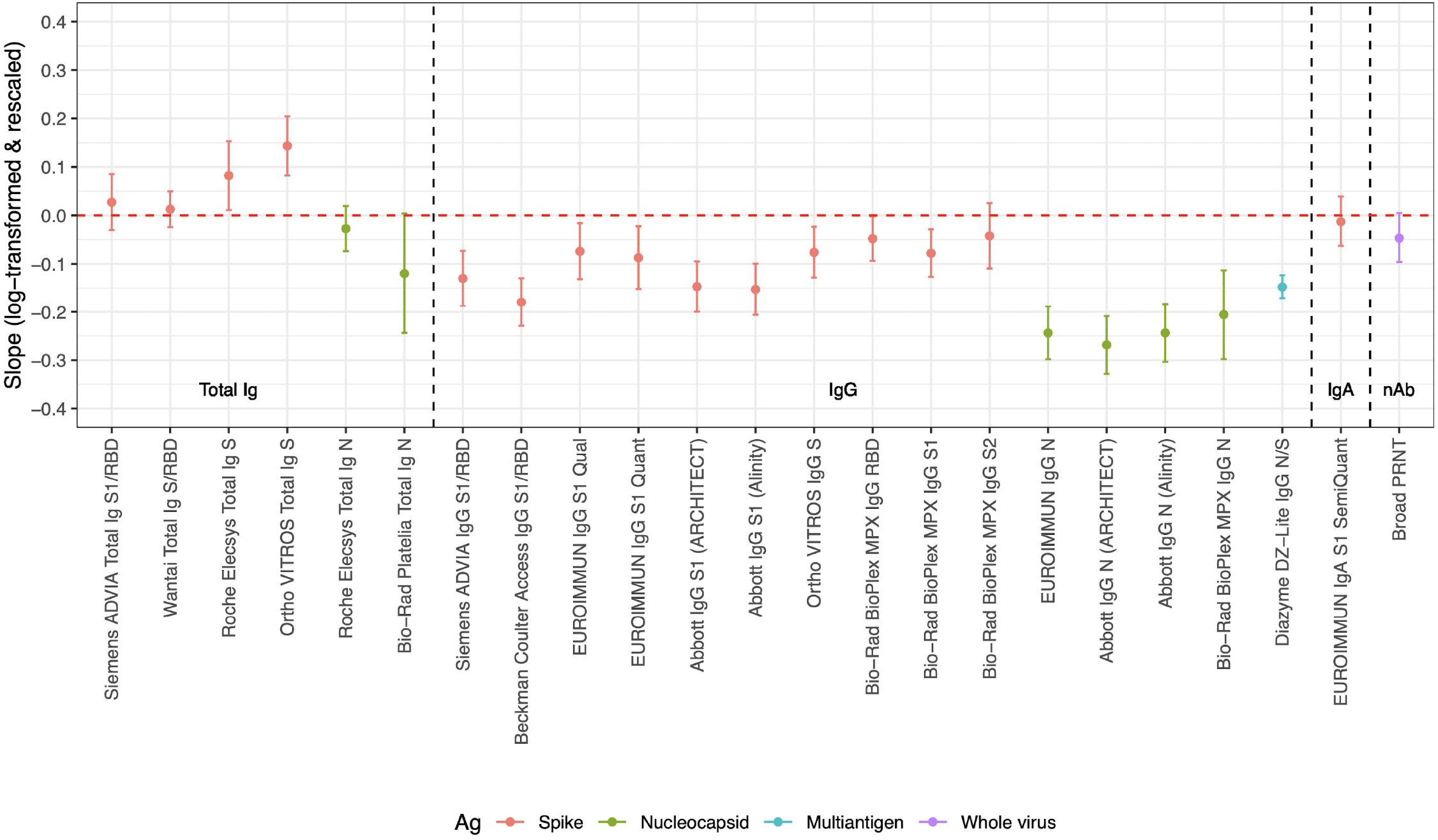
Proportion of donors with detectable SARS-CoV-2 antibodies in the longitudinal COVID-19 convalescent plasma (CCP) donor cohort with donations sorted into time bins relative to index CCP donation. Time bin labels on x axis are denoted with square brackets to indicate inclusive boundaries and round brackets to indicate exclusive boundaries. Donors that contributed more than one donation in a time bin contributed the fractional proportion reactive to the numerator and 1 to the denominator for estimation of proportion reactive in the time bin. Symbols indicate point estimates of proportion reactive and bars indicate 95% confidence intervals (Wilson score). See Table 1 for assay details. S, spike protein; RBD, receptor binding domain; N, nucleocapsid; Ag, antigen; Ab, antibody; Ig, immunoglobulin; PRNT, plaque reduction neutralization test. *Ortho VITROS Total Ig anti-S reactivity was required for qualification of continued CCP donation, and therefore shows 100% detection in all time bins by definition.

Regression models of quantitative signal intensity over time showed statistically significant declining reactivity in some assays. All anti-S total Ig (“direct” antigen sandwich format) assays showed stable or increasing reactivity, while all IgG assays showed declining reactivity over time (Figure 4, panel A). Anti-N assays showed more rapid waning than anti-S assays, with multivariable regression confirming that both assay format and antigen (Ag) target are important rate of waning predictors. Amongst assays that showed statistically significant declining reactivity, estimated half-lives varied from 41 to 574 days (median: 91 days)(Figure 4, panel B).

**Figure 4.**
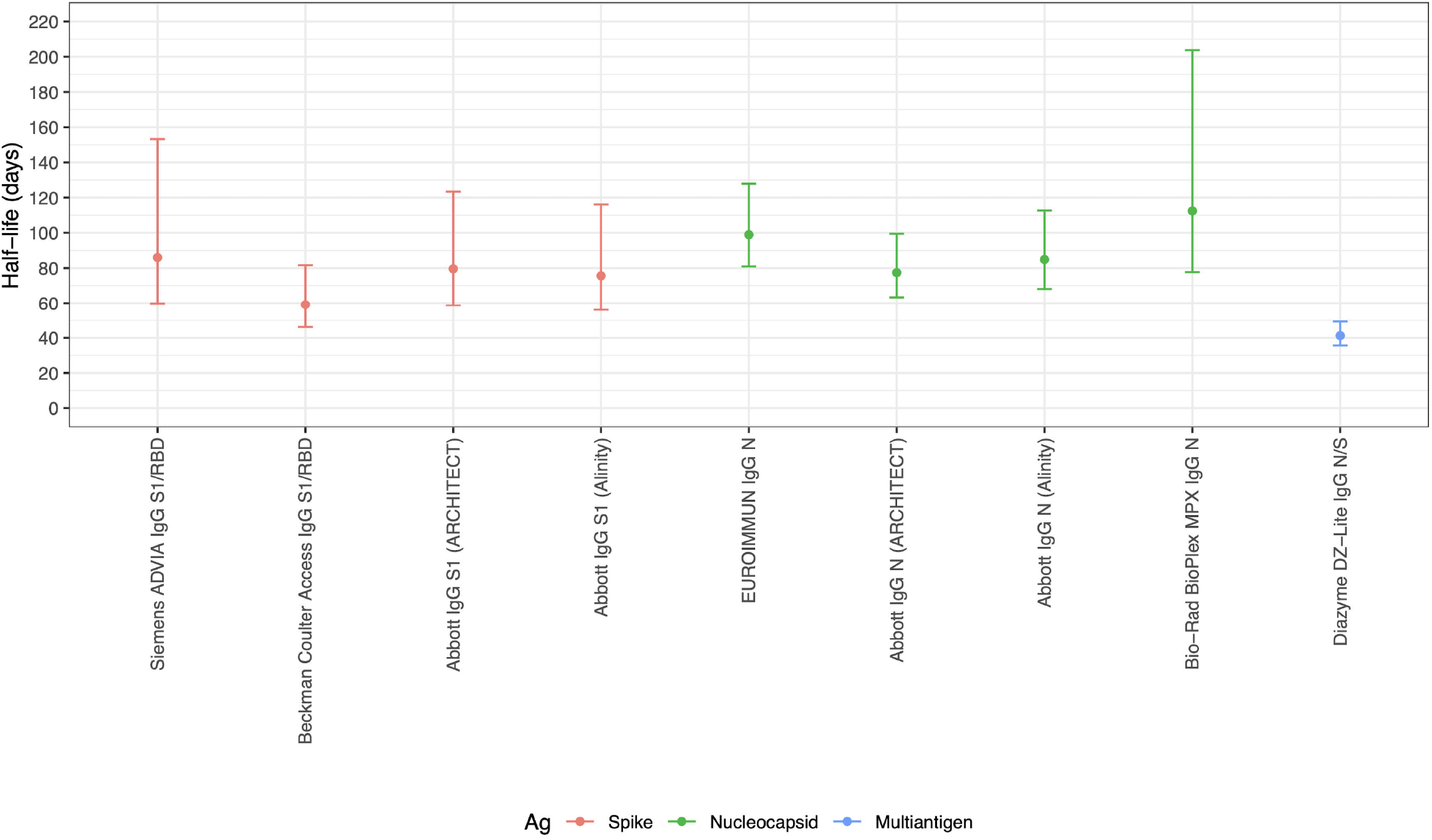
Durability of antibody detection as assessed by mixed effects regression modelling. Panel A: Average (‘fixed’) slopes from linear mixed effects regression models with donor random effects, fit to rescaled and log-transformed quantitative assay signal. Panel B: Assay signal half-lives post index donation for assays demonstrating rapid waning of seroreactivity over time (upper bound on half-life less than 220 days), estimated based on linear mixed effects regression models. S, spike protein; RBD, receptor binding domain; N, nucleocapsid; Ag, antigen; Ab, antibody; Ig, immunoglobulin. See Table 1 for assay details.

All assays included in the study showed ‘good’ or ‘excellent’ quantitative repeatability as reflected in the ICC, i.e., ICCs ≥0.75 and ≥0.9 [10], respectively, with the exception of the Wantai assay that had an ICC below 0.6 (Figure 5, Appendix Table 2). CVs were generally <10% for low and medium titer blinded replicate specimens, and somewhat higher for high titer specimens, ranging from ∼20% to over 100% (Appendix Table 3). The Ortho VITROS anti-S and Roche Elecsys anti-N total Ig assays had notably low CVs on most replicate specimens (generally below 10%).

**Figure 5.**
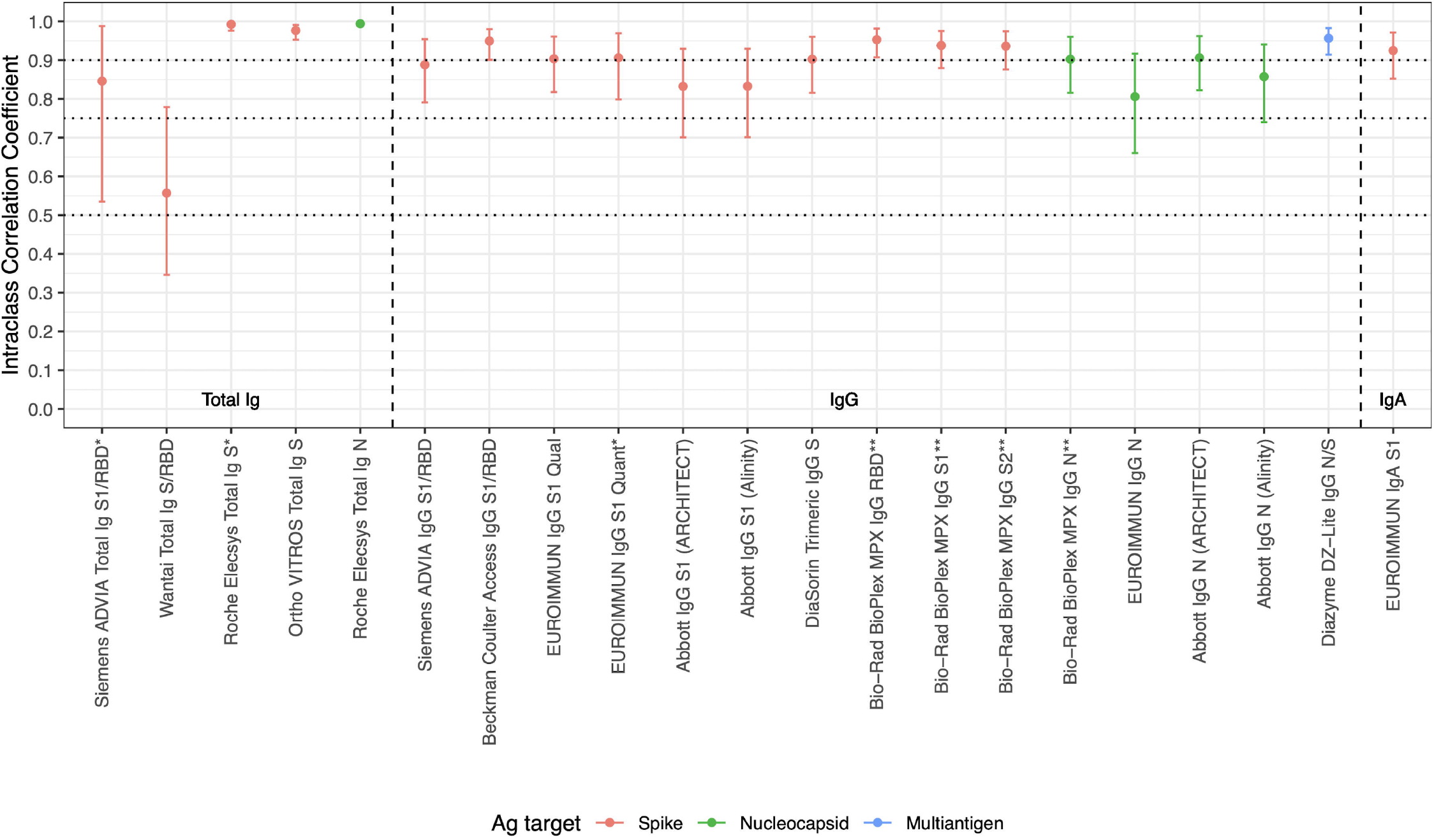
Intraclass correlation coefficients based on blinded replicate sample testing, reflecting the proportion of total variance that is between-sample rather than within-sample variability. S, spike protein; RBD, receptor binding domain; N, nucleocapsid; Ag, antigen; Ab, antibody; Ig, immunoglobulin. See Table 1 for assay details. * Results falling outside the primary measurement range excluded. ** On-board dilutions were used to estimate reactivity in specimens where initial results fell outside the primary measurement range. *Horizontal dotted lines show conventional (although arbitrary) thresholds for “moderate” (0*.*5), “good” (0*.*75) and “excellent” (0*.*9) repeatability [10]*.

Dilutional performance was generally good, with most assays demonstrating reasonable linearity in the relationship between expected and observed reactivity above the assay cutoff (Appendix Figure 5). Assays with greater dynamic ranges tended to show a linear dilutional response even below the cutoff. Most assays had a well-defined inflection point, representing a level of reactivity below which the dilutional response was not linear.

For most assays all 24 serosilent specimens were non-reactive, 7 assays had 1/24 specimen reactive, 2 assays had 2/24 specimens reactive. The vast majority of apparent serosilent specimens were not detected using any of the serological assays included in this study (Appendix Figure 6). For the single seroconversion series, most assays show seroconversion over the same two-week timeframe, providing little evidence of variable sensitivity relative to time of infection (Appendix Figure 7).

## Discussion

This study characterized 21 commercial SARS-CoV-2 serological assays, supporting the development, validation, and implementation of testing algorithms for serosurveillance programs, including algorithms that can distinguish natural infection from vaccine induced seroreactivity.

The three most critical characteristics for assays used to conduct serosurveillance are 1) sensitivity including an assay’s ability to detect antibodies following asymptomatic and mildly symptomatic infections potentially resulting in weak Ab responses [11-13], 2) specificity to minimize the impact of false positives on seroprevalence estimates, and 3) the ability to durably detect Ab responses for accurate estimation of cumulative infections. For serosurveillance in the context of widespread spike-based vaccine implementation, algorithms that combine assays with these characteristics and different Ag targets can differentiate natural infection from vaccine induced seroreactivity. The ability to detect reinfections and vaccine breakthrough infections based on development of anti-N reactivity, including those from variants,[14-18] requires quantitative assays with wide dynamic ranges, including the ability to extend the dynamic range through dilution, and good quantitative repeatability to enable detection of anamnestic boosting of humoral immune responses. However in regions where whole virus vaccines are in use, alternative algorithms should be considered.

The impact of particular performance characteristics on interpretation of serosurveillance data is context dependent. Ideal assays for serosurveillance applications (eg total Ig assays demonstrating excellent specificity, sensitivity and durability of Ab detection) may not be a viable option and other factors such as cost, logistics and regulatory process may influence assay availability and selection, particularly in resource constrained settings. However, if assay performance has been robustly characterized, statistical adjustments can be made in the estimation of seroprevalence.

It is common practice for assay manufacturers to determine sensitivity based on timing of seroconversion relative to diagnostic testing or clinical disease. Because this clinical diagnostic definition of sensitivity may not be the most relevant criterion in cases where there is a high rate of mildly symptomatic or asymptomatic infections, alternate definitions of true positivity should be considered. Thus, in this study focused on serosurveillance applications, we used multiple definitions to assess sensitivity. These definitions allowed us to assess sensitivity in practical serosurveillance contexts. Of particular note, the inclusion of all CCP donors results in lower sensitivity estimates consequent to inclusion of serosilent infection cases, whereas the requirement for neutralization activity excludes those cases resulting in higher sensitivity estimates.

The ability to detect past infections long after the resolution of symptoms is key to accurately estimate cumulative incidence of infections based on seroreactivity rates; otherwise, complex and unvalidated adjustments for seroreversion may be required[17, 19, 20]. To evaluate the durability of humoral immunity after natural infection and vaccination, and to detect anamnestic boosting of Abs following reinfection or vaccine breakthrough, it is necessary to detect both changes in quantitative signal intensity and qualitative results over time, and this requires assays with wide dynamic range and quantitative precision. Seroreactivity levels on such assays may still plateau at the upper limit of quantitation. Dilution of specimens, which many platforms can perform automatically, extend dynamic ranges enabling quantitation of high titer specimens as demonstrated by the Bio-Rad BioPlex assay. Furthermore, rates of waning immunity are difficult to assess using assays with narrow dynamic ranges that constrain detection of declining reactivity, which may persist at the upper limit of quantification. Although qualitative seroreversion was observed over the timescale evaluated in some assays including ones with narrow dynamic range (Figure 3), further studies are required to assess the durability of detection over longer timescales. Quantitation of very low-level reactivity is possible in assays demonstrating linearity of dilutional performance below the manufacturer defined thresholds for reactivity, which are generally set to maintain high specificity. Quantitation of high-level reactivity requires assays with a wide dynamic range, or testing of dilutions to extend the measurement range, which is more practical on platforms that support on-board dilutions.

We observed that all anti-S total Ig (“direct” antigen sandwich format) assays showed stable or increasing reactivity, while all but one IgG assays showed declining reactivity over time presumably due to continued maturation of Ab affinity and/or avidity resulting in increasing signal intensity in these assays [21-23]. Anti-N assays showed more rapid waning than anti-S assays, with multivariable regression confirming that both assay format and Ag target are important rate of waning predictors.

The relatively stable detection of neutralizing activity up to four months post index donation demonstrates that in the cross-sectional CCP sample set used in the sensitivity analysis, any waning of nAb titers was very unlikely to have taken place by the time specimens were collected and would therefore not have biased sensitivity analyses based on neutralizing activity.

Although there was sporadic reactivity in a few specimens from serosilent cases, most assays included in this evaluation tested non-reactive on all specimens. This corroborates the findings of other studies [11, 24] indicating that some infected individuals do not develop a detectable systemic humoral immune response to SARS-CoV-2 infection.

The best performing assays for serosurveillance applications in this evaluation were high-throughput total Ig antigen sandwich format assays, as they met the three key performance criteria of durable Ab detection, sensitivity and specificity. The Ortho and Roche total Ig assays that target S and N antibodies, performed well and are currently employed in large scale serosurveillance studies in the US, Canada, the UK and other countries, including the CDC-nationwide blood donor seroprevalence study (<u>COVID Data Tracker</u>). The Wantai assay has been widely used in serosurveillance globally [25-27]; while this demonstrated lower specificity and reproducibility than the best performing assays, it performs adequately for serosurveillance with accounting for those limitations. Several other assays, including the Abbott IgG anti-N and EUROIMMUN IgG anti-S assays, have been employed in large-scale serosurveillance, but require adjustments for rapid waning and seroreversion to estimate cumulative incidence or attack rates, especially over longer periods and multiple epidemic waves. This study provides critical data that can be applied to adjust for waning in other studies.

This study has several limitations. Asymptomatic cases are underrepresented in the panel as CCP donors qualify based on recovery from symptomatic infection, potentially resulting in overestimation of sensitivity. The assessment of durability of bAb detection is based on CCP donations from donors whose continued qualification required ongoing Ortho VITROS Total Ig anti-S1 reactivity. Although these CCP donors do not have documented dates of NAT-positivity, symptom onset or resolution, the first donations were generally within 1-2 months of symptom resolution [5]. To address these limitations we developed approaches to adequately characterize sensitivity and durability of reactivity. The number of specimens included in the dilutional series subpanels are not sufficient for robust assessment of endpoint dilutional sensitivity.

This study provides a standardized, comparative assessment of 21 SARS-CoV-2 Ab assays from major commercial manufacturers and allows for identification of optimal assays and testing algorithms for serosurveillance applications in various contexts. These results also provide performance data applicable to other serological testing use cases relevant to clinicians, public health organizations, laboratorians, and emergency response planners.

## Data Availability

The analytic data set is available upon request.

## Acknowledgements

We thank the manufacturers for participation and/or provision of reagents: Ortho Clinical Diagnostics, Euroimmun, Roche Diagnostics, Diasorin, Siemens, Abbott, Bio-Rad, Quotient, Diazyme, Beckman Coulter, Wantai; the VRI Research Operations Core and Core Immunology Labs, and the testing laboratories.

