## Supplemental Material for "Evaluation of commercially available high-throughput SARS-CoV-2 serological assays for serosurveillance and related applications"

Running title: Evaluation of high-throughput COVID-19 serology assays

Mars Stone* ^1,2^, Eduard Grebe* ^1,2,3^, Hasan Sulaeman^1^, Clara Di Germanio^1^, Honey Dave^1^, Kathleen Kelly^1^, Brad Biggerstaff^4^, Bridgit O. Crews^5^, Nam Tran^6^, Keith R. Jerome^7^, Thomas N. Denny^8^, Boris Hogema^9^, Mark Destree^10^, Jefferson M. Jones^11^, Natalie Thornburg^11^, Mel Krajden^12^, Graham Simmons^1,2^, Steve Kleinman^13^, Larry J. Dumont^1,14^, Michael P. Busch^1,2^

Author affiliations:

Vitalant Research Institute, San Francisco CA, USA (M. Stone, E. Grebe, H. Sulaeman, C, Di Germanio, H. Dave, K. Kelly, G. Simmons, L.J. Dumont, M.P. Busch); Department of Laboratory Medicine, University of California, San Francisco, CA, USA (M. Stone, E. Grebe, G. Simmons, M.P. Busch); SACEMA, Stellenbosch University, Stellenbosch, South Africa (E. Grebe); Centers for Disease Control and Prevention, Fort Collins, CO, USA (B. Biggerstaff)

University of California Irvine Medical Center, Orange, CA, USA (B.O. Crews)

Department of Pathology and Laboratory Medicine University of California, Davis, CA, USA (N. Tran); Fred Hutchinson Cancer Research Center, University of Washington, Seattle, WA, USA (K.R. Jerome); Duke Human Vaccine Institute, Duke University, Durham, NC, USA (T. Denny); Department of Blood-Borne Infections, Sanquin Research, Amsterdam, The Netherlands (B. Hogema); BloodWorks North West, Seattle, WA, USA (M. Destree)

Centers for Disease Control and Prevention, COVID-19 Response Team, Atlanta GA, USA (J.M. Jones, N. Thornburg); British Columbia Centre for Disease Control, Vancouver, Canada (M. Krajden); Department of Pathology and Laboratory Medicine, University of British Columbia, Vancouver, Canada (S. Kleinman); University of Colorado School of Medicine, Denver, CO, USA (L.J. Dumont).

* These first authors contributed equally to this article.

Disclaimer: The findings and conclusions in this report are those of the authors and do not necessarily represent the official position of the Centers for Disease Control and Prevention.

Word count: 3515/3500

Abstract 150/150

Keywords: COVID-19, serosurveillance, serology, assay

Corresponding author: Mars Stone, PhD., Vitalant Research Institute, San Francisco CA, USA, Department of Laboratory Medicine, University of California San Francisco, CA, USA. 270 Masonic Ave, San Francisco, CA 94118. Phone (415) 354-1389, fax (415) 775-3859.

**Methods**

*Assay selection, panel development and testing*

Key characteristics of the assays included, such as format and configuration, antigen composition, and immunoglobulin target, are summarized in Table 1. Uniquely blinded panels were distributed to manufacturer-approved testing laboratories. Compiled data were analyzed and used to determine performance characteristics (sensitivity, specificity, repeatability, dilutional performance, durability of reactivity over time, assay concordance, and correlations of levels of observed reactivity with neutralizing antibody [nAb] titers).

**Discussion**

This study characterized 21 commercial SARS-CoV-2 serological assays. This evaluation and these data can support the development, validation, and implementation of testing algorithms for serosurveillance programs, including algorithms that can distinguish natural infection from vaccine induced seroreactivity. This study demonstrated that the signal intensity of several assays had wide dynamic ranges, excellent repeatability, and high precision, suggesting that signal intensity values can be quantitated (e.g., followed longitudinally). This enables using the assays to detect waning immunity, reinfections, and vaccine-breakthrough infections.

With rapid implementation of SARS-CoV-2 vaccination, serologic testing algorithms must be carefully considered. In settings where spike-based vaccines are in use, combinations of S- and N-based assays or multiplexed S/N assaysthat detect and differentiate vaccine from natural infection induced seroreactivity, ideally on the same platform either sequentially or in parallel, are needed to accurately interpret seroreactivity rates, monitor vaccine penetrance as well as to detect vaccine breakthrough infections based on development of anti-N reactivity. This is of increasing concern given the recent evolution and spread of SARS-CoV-2 variants [16]. The algorithm adopted for the CDC-nationwide blood donor seroprevalence study ([COVID Data Tracker](https://covid.cdc.gov/covid-data-tracker/#cases_casesper100klast7days)) uses S-based screening and N-based confirmation to discriminate natural infection from vaccine induced seroreactivity and has the potential to monitor for breakthrough infections in vaccinated repeat blood donors. Our assessment demonstrates that a combination of assays with stable Ab detection (such as total-Ig antigen-sandwich format assays) that detect anti-S and anti-N antibodies is optimal for serosurveillance in settings where all vaccines are based on the S protein. However in regions where whole virus vaccines are in use, alternative algorithms should be considered.

The best performing assays for serosurveillance applications in this evaluation were high-throughput total Ig antigen sandwich format assays, as they met the three key performance criteria of durable Ab detection, sensitivity and specificity. The Ortho and Roche total Ig assays that target S and N antibodies, performed well and are currently employed in large scale serosurveillacne studies in the US, Canada, the UK and other countries. The Wantai assay has been widely used in serosurveillance globally [25-27]; while this demonstrated lower specificity and reproducibility than the best performing assays in this total Ig assay category, its performance is adequate for serosurveillance with robust characterization adjustments for those limitations made in deriving seroprevalence estimates.

Assays with poor quantitative repeatability are not suitable to quantify bAb titers or make inferences about immunity. In general, we recommend the use of assays with ICCs above 0.9 for quantitation. Several other assays, including the Abbott IgG anti-N and EUROIMMUN IgG anti-S assays, have been employed in large-scale serosurveillance, but require adjustments for rapid waning and seroreversion to estimate cumulative incidence or attack rates, especially over longer periods and multiple epidemic waves. This study provides critical data that can be applied to adjust for waning in other studies.

This study has several limitations. For the single seroconversion series, most assays show seroconversion over the same two-week timeframe, providing little evidence of variable sensitivity relative to time of infection (Appendix Figure 7). However, the use of a single seroconversion panel without a documented date of NAT-positivity or symptom onset/resolution is a major limitation of this analysis. Other limitations include that specimens from previously-infected individuals are limited to qualified CCP donors who recovered from symptomatic COVID-19. Asymptomatic cases are therefore underrepresented in the panel, potentially resulting in overestimation of the sensitivity of the indirect Ig tests. The assessment of durability of bAb detection is limited to longitudinal CCP donations from donors who remained reactive on the Ortho VITROS Total Ig anti-S1 assay at all timepoints to continue qualification for CCP donations. Although these CCP donors do not have documented dates of NAT-positivity, symptom onset or resolution, the first donations were generally within 1-2 months of symptom resolution [5]. Peak reactivity for estimating time to seroreversion cannot be defined and this complicates interpretation of slopes in reactivity over time. To address these limitations multiple analyses using distinct approaches were conducted to characterize sensitivity and durability of reactivity. The number of specimens included in the dilutional series and in the serosilent case subpanels are not sufficient for robust assessment of endpoint dilutional sensitivity and ability to detect prior infections in individuals who do not seroconvert.

This study provides a standardized, comparative assessment of 21 SARS-CoV-2 Ab assays from major commercial manufacturers and allows for identification of optimal assays and testing algorithms for serosurveillance applications in various contexts. The results of the study also provide performance data relevant to other serological testing use cases. While the present analysis is focused on the serosurveillance use case, these data are also relevant to clinicians, public health organizations, laboratorians, and emergency response planners. Further studies are in progress using these data with additional panels including vaccinated donors, to develop and apply testing algorithms for detection and characterization of immune persistence, vaccine responses in naïve and previously infected individuals, detection of reinfections and vaccine breakthrough infections, as well as characterization of the potency of passive immunotherapies.

Acknowledgements: We thank the manufacturers for participation and/or provision of reagents: Ortho Clinical Diagnostics, Euroimmun, Roche Diagnostics, Diasorin, Siemens, Abbott, Bio-Rad, Quotient, Diazyme, Beckman Coulter, Wantai; the VRI Research Operations Core and Core Immunology Labs, and the testing laboratories.

**Funding source:** This work was supported by research contracts from the Centers for Disease Control and Prevention (CDC Contract 75D30120C08170).

**Tables and Figures**

| **Manufacturer** | **Assay** | **Ig Target** | **Antigen** | **Assay Format** | **Reported units** | **Testing Lab** |
| --- | --- | --- | --- | --- | --- | --- |
| **Ortho** | VITROS Immunodiagnostic Products Anti-SARS-CoV-2 Total Ig | Total Ig | S1 | Double-antigen sandwich CLIA | S/CO | Vitalant Research Institute |
|  | VITROS Immunodiagnostic Products Anti-SARS-CoV-2 IgG | IgG | S1 | Double-antigen sandwich CLIA | S/CO | CTS |
| **EUROIMMUN** | Anti-SARS-CoV-2 NCP ELISA | IgG | N | Indirect IgG EIA | S/CO | Advent Health |
|  | Anti-SARS-CoV-2 ELISA | IgG | S1 | Antigen sandwich ELISA | S/CO |  |
|  | Anti-SARS-CoV-2 QuantiVac ELISA | IgG | S1 | Antigen sandwich ELISA | RU/ml |  |
|  | Anti-SARS-CoV-2 ELISA | IgA | S1 | Antigen sandwich ELISA | S/CO |  |
| **Roche** | Elecsys Anti-SARS-CoV-2 N on cobas | Total Ig | N | Double-antigen sandwich CLIA | COI | University of California, Davis |
|  | Elecsys Anti-SARS-CoV-2 S on cobas | Total Ig | S1/S2/RBD | Double-antigen sandwich CLIA | U/ml |  |
| **DiaSorin** | LIAISON 28 SARS-CoV-2 TrimericS IgG | IgG | TrimericS | IgG magnetic particle CLIA | AU/ml | British Colombia Centers for Disease Control and Prevention |
| **Siemens** | ADVIA Centaur SARS-CoV‑2 Total Ig | Total Ig | S1/RBD | Ag sandwich CLIA | S/CO |  |
|  | ADVIA Centaur SARS-CoV‑2 IgG | IgG | S1/RBD | Ag sandwich CLIA | Index |  |
| **Abbott** | SARS-CoV-2 IgG N on ARCHITECT | IgG | N | CMIA | AU/ml | Duke Human Vaccine institute |
|  | SARS-CoV-2 IgG S1 on ARCHITECT | IgG | S1 | CMIA | S/CO |  |
|  | SARS-CoV-2 IgG N on Alinity | IgG | N | CMIA | S/CO | Fred Hutchinson Cancer Research Center |
|  | SARS-CoV-2 IgG S1 on Alinity | IgG | S1 | CMIA | AU/ml |  |
| **Bio-Rad** | Platelia SARS-CoV-2 Total Ab (Evolis) | Total Ig | N | One-step antigen capture | S/CO | BloodWorks NorthWest |
|  | BioPlex 2200 SARS-CoV-2 IgG Panel | IgG | RBD, S1, S2, N | Multiplexed microbeads two-step assay | S/CO |  |
| **Quotient** | MosaiQ™ COVID-19 Antibody Microarray | IgM/IgG | S1/S2 | Array | Qual only |  |
| **Diazyme** | DZ-Lite SARS CoV-2 | IgG | N & S1/S2 | IgG microbead CLIA | S/CO | University of California, Irvine |
| **Beckman Coulter** | Access SARS-CoV-2 IgG | IgG | S1 RBD | IgG 2-step paramagnetic particle CLIA | S/CO |  |
| **Wantai** | SARS-CoV-2 Total Ig | Total Ig | S1 RBD | Total Ig sandwich ELISA | S/CO | Sanquin |

**Table 1.** Serological assays and key characteristics. S, spike protein; RBD, receptor binding domain; N, nucleocapsid; Ag, antigen; Ab, antibody; Ig, immunoglobulin; S/CO, signal to cutoff ratio; EIA, enzyme immunoassay; ELISA, enzyme-linked immunosorbent assay;RU, relative units, AU, arbitrary units; CLIA, Chemiluminescent immunoassay; CMIA, Chemiluminescent microparticle immunoassay.

Current US regulatory status can be obtained on the FDA website at:

<https://www.fda.gov/medical-devices/coronavirus-disease-2019-covid-19-emergency-use-authorizations-medical-devices/in-vitro-diagnostics-euas-serology-and-other-adaptive-immune-response-tests-sars-cov-2> .

| **Group** | **Description** | **Number of specimens** |
| --- | --- | --- |
| **Sensitivity sub-panel** |  |  |
|  | 130 cross sectional CCP donor plasma specimens. | 185 |
| **Specificity sub-panel** |  |  |
|  | Pre-pandemic blood donor specimens collected prior to 2020 and demonstrated  to be anti-SARS-CoV-2 negative by RVP neutralization testing. | 459 |
| **Ab persistence sub-panel** |  |  |
|  | Longitudinal specimens from 26 donors with at ≥4 CCP donations 84-150 days post index donation. | 209 |
| **Seroconversion sub-panel** |  |  |
|  | Longitudinal specimens from a single source plasma donor with acute SARS-CoV-2 infection | 14 |
| **Dilutional Performance sub-panel** |  |  |
|  | Serial dilutions of five specimens from Sensitivity sub-panel; neat (6 replicates), 1:40, 1:80, 1:160, 1:320, 1:640 analogous to nAb testing | 55 |
| **Serosilent cases** |  |  |
|  | Individual CCP donors non-reactive by S and N anti-SARS-CoV-2 Total Ig | 27 |
| **Repeatability sub-panel** |  |  |
|  | Six blinded replicates each of 15 CCP specimens | 90 |

Current US regulatory status can be obtained on the FDA website at:

<https://www.fda.gov/medical-devices/coronavirus-disease-2019-covid-19-emergency-use-authorizations-medical-devices/in-vitro-diagnostics-euas-serology-and-other-adaptive-immune-response-tests-sars-cov-2>

**Table 2.** Composition of the assessment panel. CCP, COVID convalescent plasma, S, spike protein; RBD, receptor binding domain; N, nucleocapsid; Ag, antigen; Ab, antibody; Ig, immunoglobulin; RVP, reporter viral particle; bAb, neutralizing antibodies.

**A**

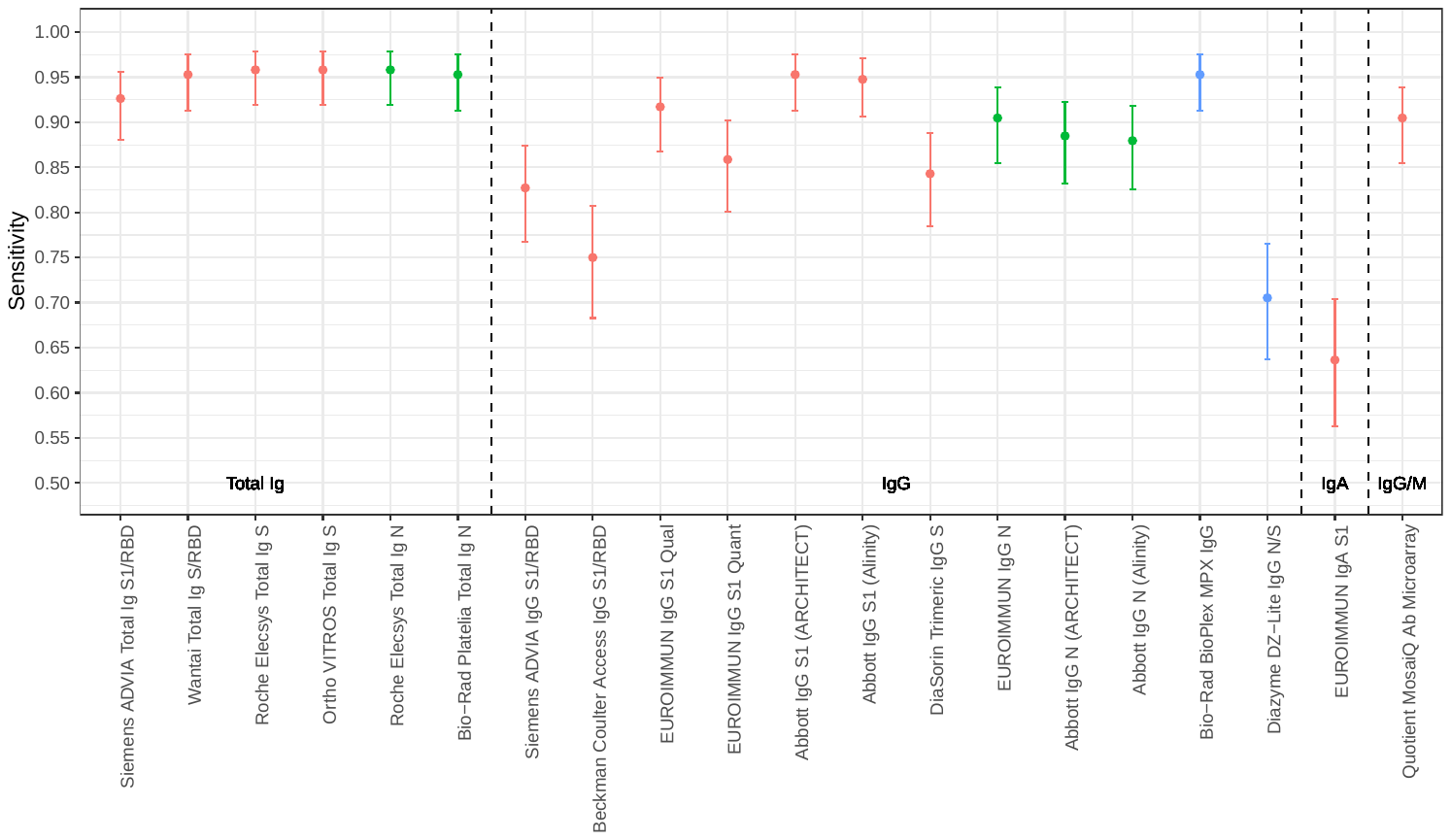

**B**

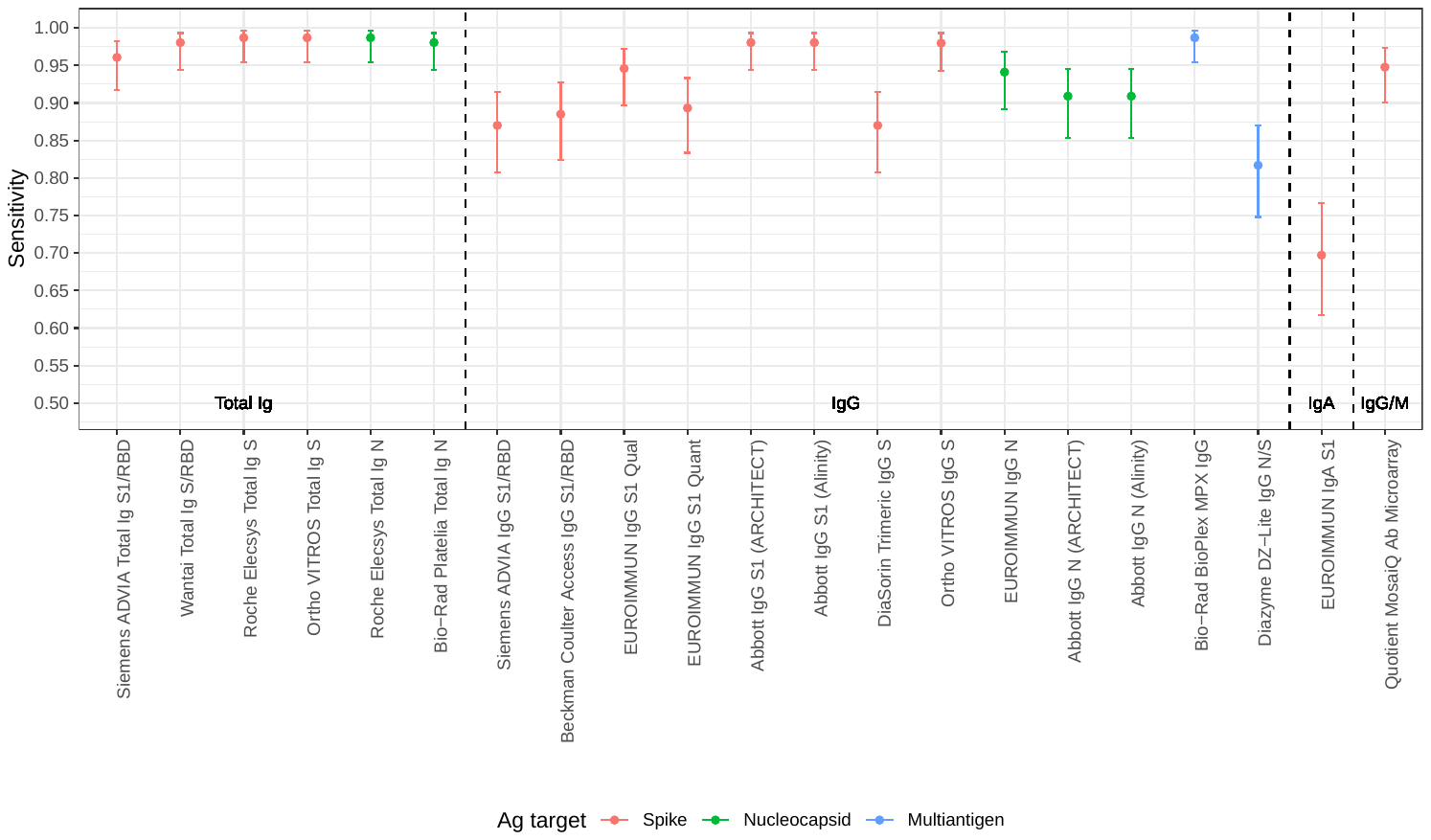

**C**
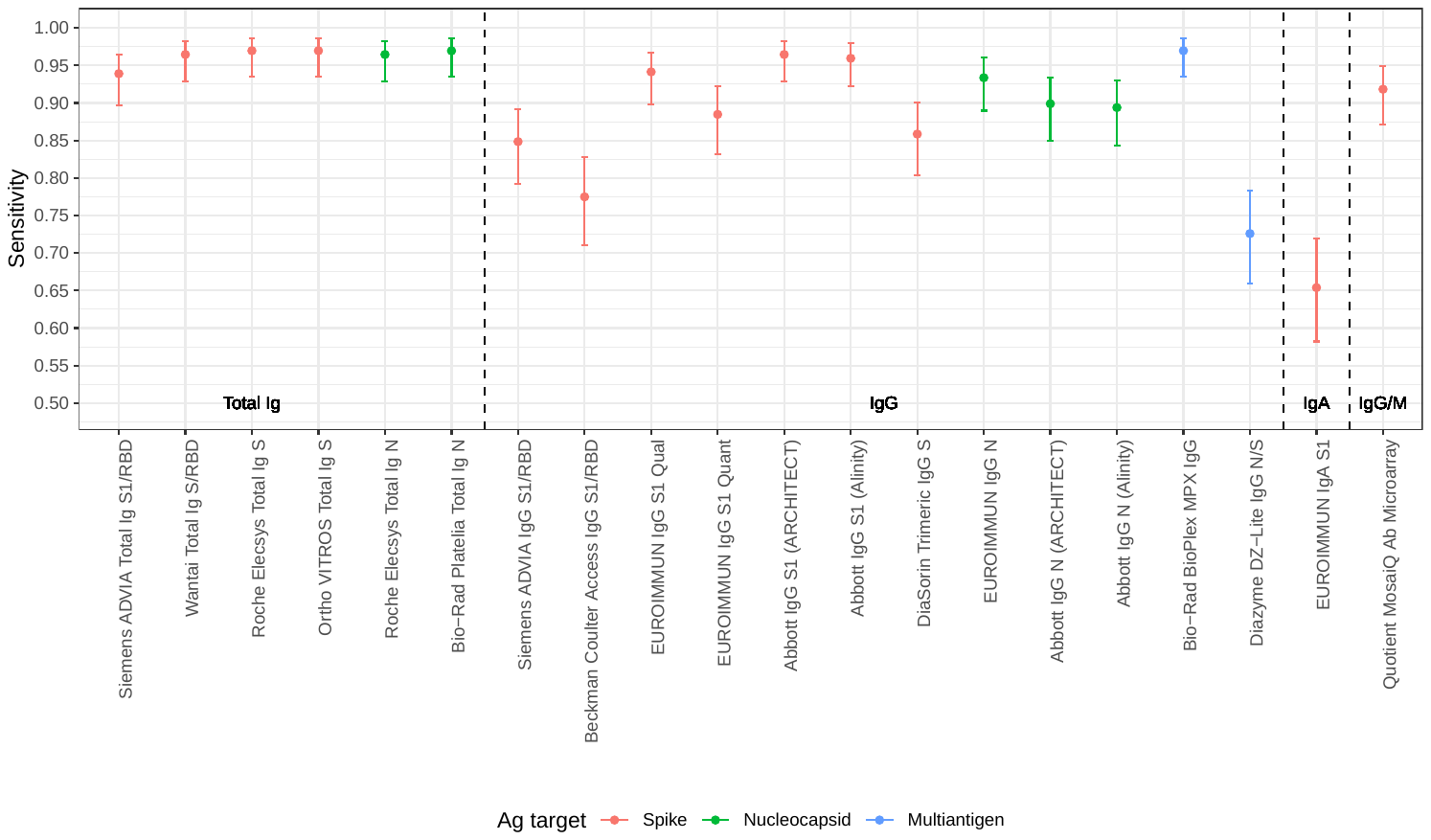

**Figure 1.** Sensitivity of SARS-CoV-2 serological assays using three definitions of a “true positive.”

Panel A: Positivity defined by qualification as CCP donor (excluding purposely selected serosilents)

Panel B: Positivity defined by neutralizing activity measured by Broad PRNT

Panel C: Positivity defined by ‘operational standard’ (3 or more bAb assays reactive).

Dots indicate point estimates and bars indicate Wilson score 95% confidence intervals.

*The Ortho VITROS IgG S assay is included only in panel B, because the assay required use of serum for testing, thus only* *specimens with available serum and neutralizing data were tested .* S, spike protein; RBD, receptor binding domain; N, nucleocapsid; Ag, antigen; Ab, antibody; Ig, immunoglobulin; PRNT, plaque reduction neutralization test. See Table 1 for assay details.

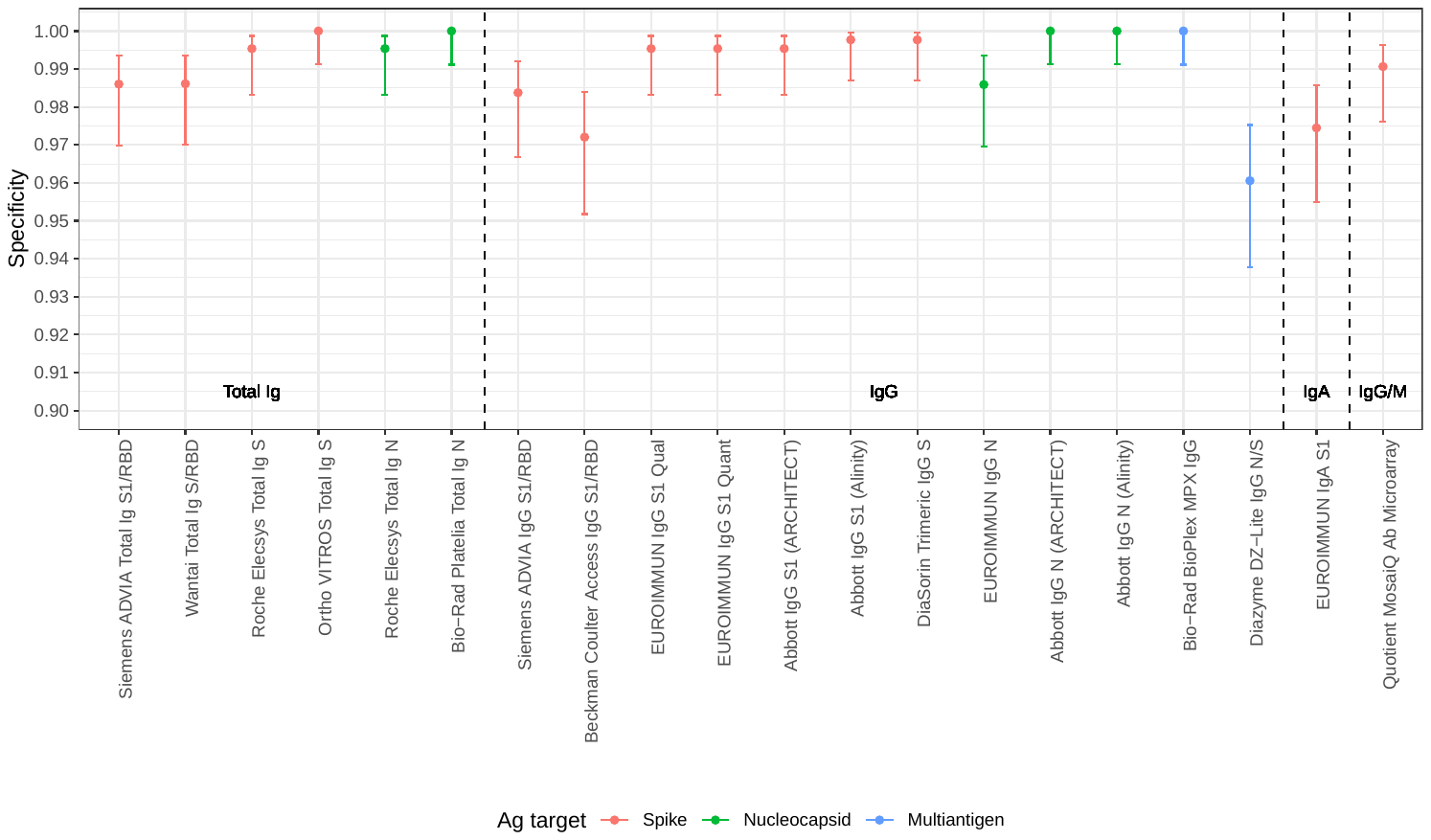

**Figure 2.** Specificity of SARS-CoV-2 serological assays in pre-COVID-19 negative control specimens.*

*Dots indicate point estimates and bars indicate Wilson score 95 confidence intervals. S, spike protein; RBD, receptor binding domain; N, nucleocapsid; Ag, antigen; Ab, antibody; Ig, immunoglobulin. See Table 1 for assay details.

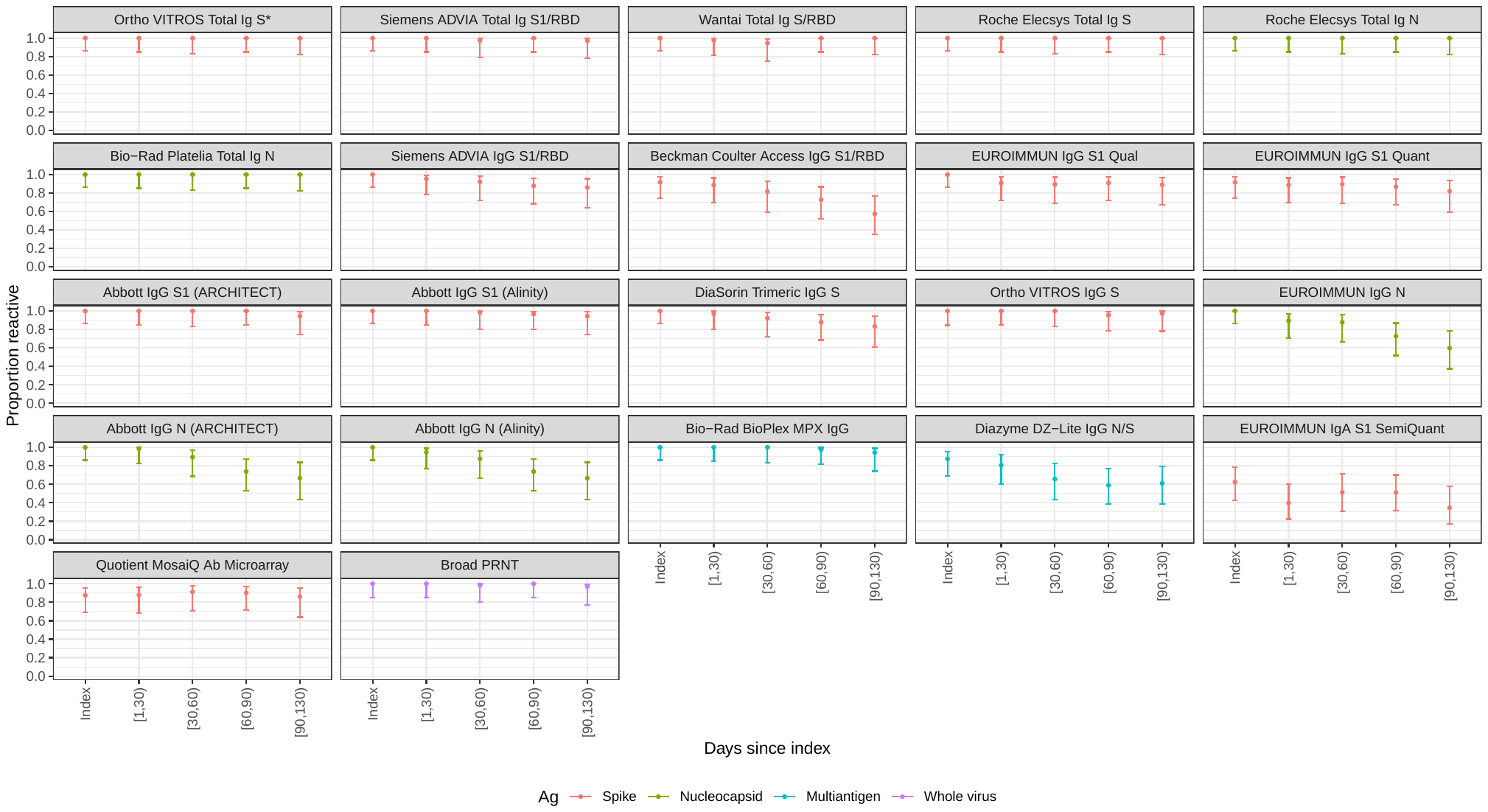

**Figure 3:** Proportion of donors with detectable SARS-CoV-2 antibodies in the longitudinal COVID-19 convalescent plasma (CCP) donor cohort with donations sorted into time bins relative to index CCP donation. Time bin labels on x axis are denoted with square brackets to indicate inclusive boundaries and round brackets to indicate exclusive boundaries. Donors that contributed more than one donation in a time bin contributed the fractional proportion reactive to the numerator and 1 to the denominator for estimation of proportion reactive in the time bin. Symbols indicate point estimates of proportion reactive and bars indicate 95% confidence intervals (Wilson score). See Table 1 for assay details. S, spike protein; RBD, receptor binding domain; N, nucleocapsid; Ag, antigen; Ab, antibody; Ig, immunoglobulin; PRNT, plaque reduction neutralization test.

*Ortho VITROS Total Ig anti-S reactivity was required for qualification of continued CCP donation, and therefore shows 100% detection in all time bins by definition.

**A**

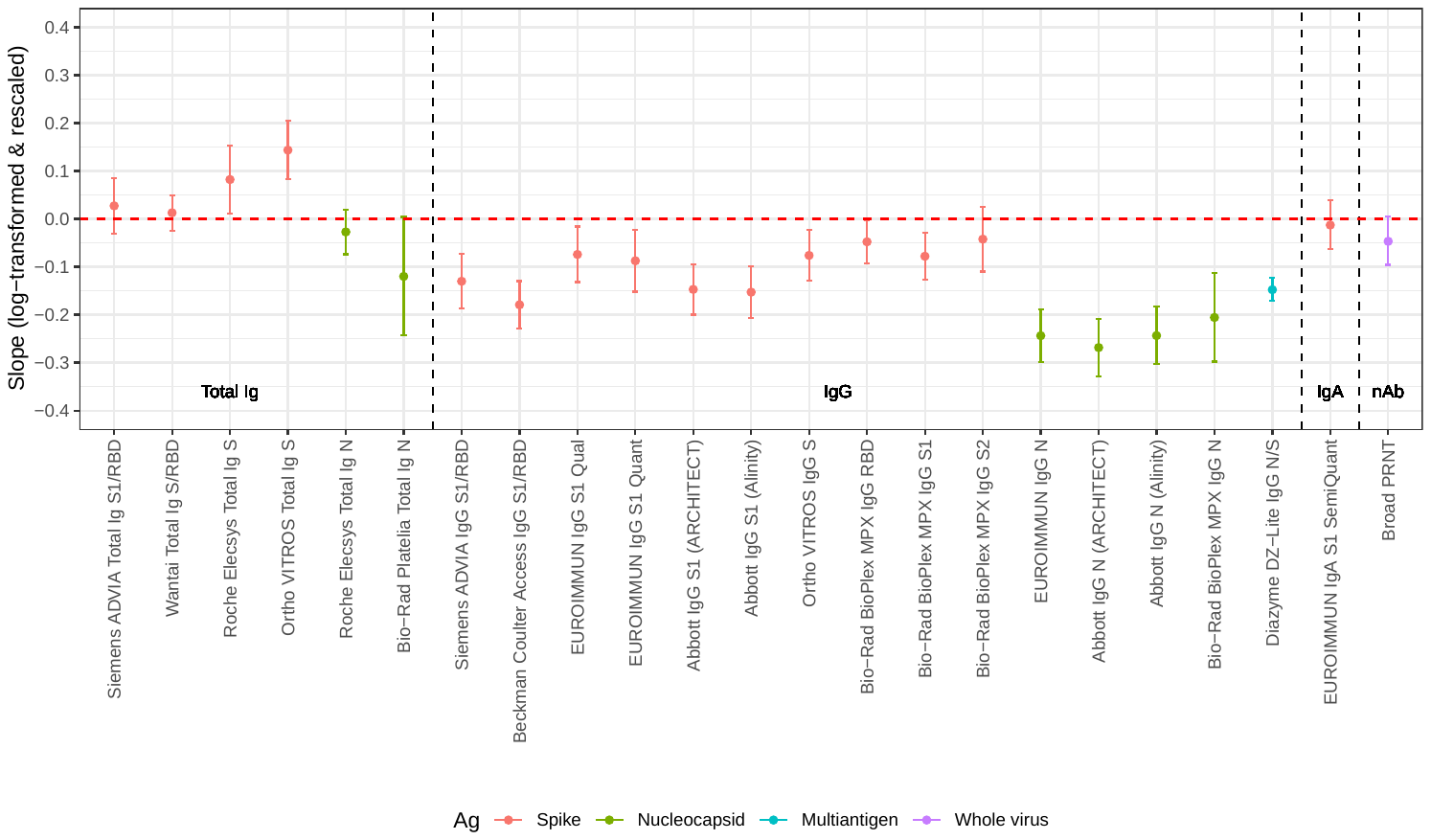

**B**

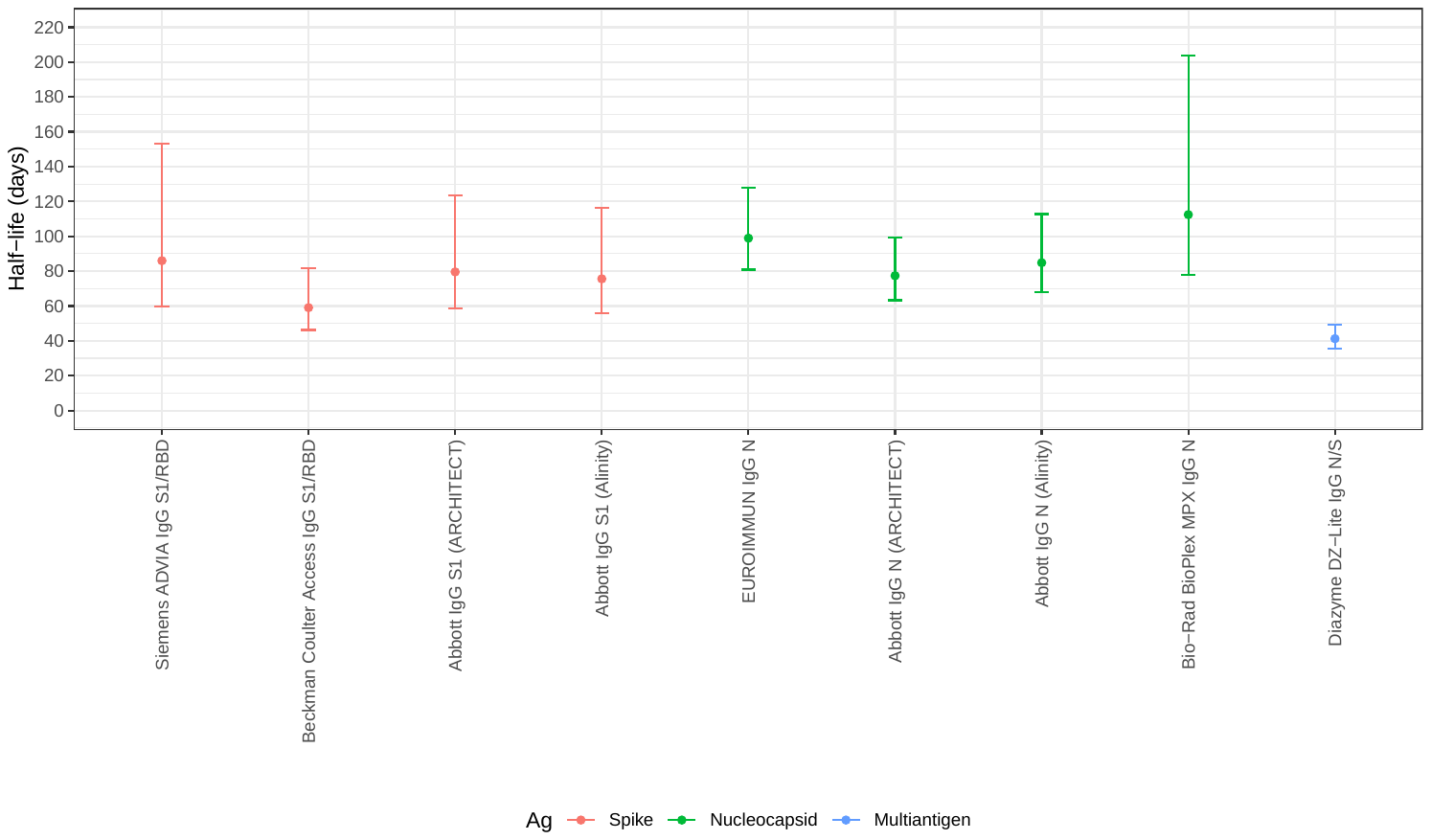

**Figure 4.** Durability of antibody detection as assessed by mixed effects regression modelling.

Panel A: Average (‘fixed’) slopes from linear mixed effects regression models with donor random effects, fit to rescaled and log-transformed quantitative assay signal.

Panel B: Assay signal half-lives post index donation for assays demonstrating rapid waning of seroreactivity over time (upper bound on half-life less than 220 days), estimated based on linear mixed effects regression models.

S, spike protein; RBD, receptor binding domain; N, nucleocapsid; Ag, antigen; Ab, antibody; Ig, immunoglobulin. See Table 1 for assay details.

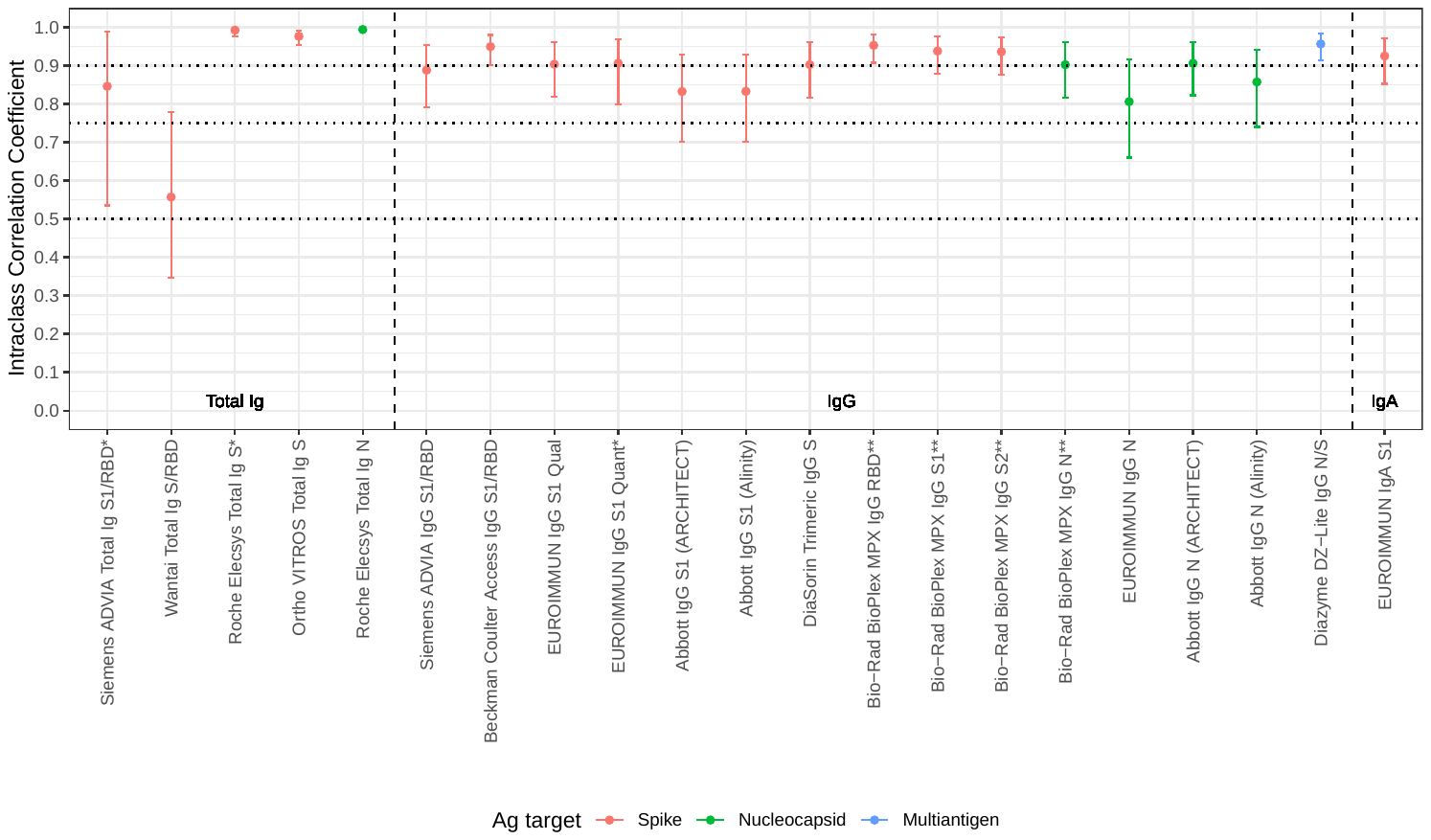

**Figure 5.** Intraclass correlation coefficients based on blinded replicate sample testing, reflecting the proportion of total variance that is between-sample rather than within-sample variability. S, spike protein; RBD, receptor binding domain; N, nucleocapsid; Ag, antigen; Ab, antibody; Ig, immunoglobulin. See Table 1 for assay details.

* Results falling outside the primary measurement range excluded. ** On-board dilutions were used to estimate reactivity in specimens where initial results fell outside the primary measurement range. *Horizontal dotted lines show conventional (although arbitrary) thresholds for “moderate” (0.5), “good” (0.75) and “excellent” (0.9) repeatability [10].*

**Supplemental Materials**

| **Assay** | **Format** | **Ag target** | **Sensitivity (definition 1)**  **(95% CI)** | **Sensitivity (definition 2)**  **(95% CI)** | **Sensitivity (definition 3)**  **(95% CI)** | **Specificity**  **(95% CI)** |
| --- | --- | --- | --- | --- | --- | --- |
| Siemens ADVIA Total Ig S1/RBD | Total Ig | Spike | 92.6% (88.0%,95.6%) | 96.1% (91.7%,98.2%) | 93.9% (89.7%,96.5%) | 98.6% (97.0%,99.4%) |
| Wantai Total Ig S/RBD | Total Ig | Spike | 95.3% (91.3%,97.5%) | 98.1% (94.4%,99.3%) | 96.5% (92.9%,98.3%) | 98.6% (97.0%,99.4%) |
| Roche Elecsys Total Ig S | Total Ig | Spike | 95.8% (92.0%,97.9%) | 98.7% (95.4%,99.6%) | 97.0% (93.5%,98.6%) | 99.5% (98.3%,99.9%) |
| Ortho VITROS Total Ig S | Total Ig | Spike | 95.8% (92.0%,97.9%) | 98.7% (95.4%,99.6%) | 97.0% (93.5%,98.6%) | 100.0% (99.1%,100.0%) |
| Roche Elecsys Total Ig N | Total Ig | Nucleocapsid | 95.8% (92.0%,97.9%) | 98.7% (95.4%,99.6%) | 96.5% (92.9%,98.3%) | 99.5% (98.3%,99.9%) |
| Bio-Rad Platelia Total Ig N | Total Ig | Nucleocapsid | 95.3% (91.3%,97.5%) | 98.1% (94.4%,99.3%) | 97.0% (93.5%,98.6%) | 100.0% (99.1%,100.0%) |
| Siemens ADVIA IgG S1/RBD | IgG | Spike | 82.7% (76.7%,87.4%) | 87.0% (80.8%,91.4%) | 84.8% (79.2%,89.2%) | 98.4% (96.7%,99.2%) |
| Beckman Coulter Access IgG S1/RBD | IgG | Spike | 75.0% (68.3%,80.7%) | 88.5% (82.4%,92.7%) | 77.5% (71.1%,82.8%) | 97.2% (95.2%,98.4%) |
| EUROIMMUN IgG S1 Qual | IgG | Spike | 91.7% (86.8%,94.9%) | 94.6% (89.7%,97.2%) | 94.1% (89.8%,96.7%) | 99.5% (98.3%,99.9%) |
| EUROIMMUN IgG S1 Quant | IgG | Spike | 85.9% (80.1%,90.2%) | 89.3% (83.4%,93.3%) | 88.5% (83.2%,92.3%) | 99.5% (98.3%,99.9%) |
| Abbott IgG S1 (ARCHITECT) | IgG | Spike | 95.3% (91.3%,97.5%) | 98.1% (94.4%,99.3%) | 96.5% (92.9%,98.3%) | 99.5% (98.3%,99.9%) |
| Abbott IgG S1 (Alinity) | IgG | Spike | 94.8% (90.6%,97.1%) | 98.1% (94.4%,99.3%) | 96.0% (92.2%,97.9%) | 99.8% (98.7%,100.0%) |
| DiaSorin Trimeric IgG S | IgG | Spike | 84.3% (78.5%,88.8%) | 87.0% (80.8%,91.4%) | 85.9% (80.3%,90.0%) | 99.8% (98.7%,100.0%) |
| Ortho VITROS IgG S | IgG | Spike | ** | 98.0% (94.2%,99.3%) | ** | ** |
| EUROIMMUN IgG N | IgG | Nucleocapsid | 90.5% (85.4%,93.9%) | 94.1% (89.2%,96.9%) | 93.4% (89.0%,96.1%) | 98.6% (97.0%,99.4%) |
| Abbott IgG N (ARCHITECT) | IgG | Nucleocapsid | 88.5% (83.2%,92.3%) | 90.9% (85.3%,94.5%) | 89.9% (84.9%,93.4%) | 100.0% (99.1%,100.0%) |
| Abbott IgG N (Alinity) | IgG | Nucleocapsid | 88.0% (82.6%,91.8%) | 90.9% (85.3%,94.5%) | 89.4% (84.3%,93.0%) | 100.0% (99.1%,100.0%) |
| Bio-Rad BioPlex MPX IgG | IgG | Multiantigen | 95.3% (91.3%,97.5%) | 98.7% (95.4%,99.6%) | 97.0% (93.5%,98.6%) | 100.0% (99.1%,100.0%) |
| Diazyme DZ-Lite IgG N/S | IgG | Multiantigen | 70.5% (63.7%,76.6%) | 81.7% (74.8%,87.0%) | 72.6% (66.0%,78.3%) | 96.1% (93.8%,97.5%) |
| EUROIMMUN IgA S1 | IgA | Spike | 63.6% (56.3%,70.4%) | 69.7% (61.7%,76.7%) | 65.4% (58.2%,71.9%) | 97.4% (95.5%,98.6%) |
| Quotient MosaiQ Ab Microarray | IgG/IgM | Spike | 90.5% (85.4%,93.9%) | 94.8% (90.0%,97.3%) | 91.8% (87.2%,94.9%) | 99.1% (97.6%,99.6%) |

**Appendix Table 1:** Sensitivity of SARS-CoV-2 serological assays using three reference definitions, and specificity in pre-COVID-19 negative control specimens. S, spike protein; RBD, receptor binding domain; N, nucleocapsid; Ag, antigen; Ab, antibody; Ig, immunoglobulin. See Table 1 for assay details.

Sensitivity definition 1: Positivity defined by qualification as CCP donor (excluding purposely selected serosilents)

Sensitivity definition 2: Positivity defined by neutralizing activity measured by Broad PRNT

Sensitivity definition 3: Positivity defined by ‘operational standard’ (3 or more bAb assays reactive).

*** Relevant specimen set not tested.*

| **Assay** | **Format** | **Ag target** | **Intraclass Correlation Coefficient**  **(95% CI)** | **N** | **k** |
| --- | --- | --- | --- | --- | --- |
| Siemens ADVIA Total Ig S1/RBD* | Total Ig | Spike | 0.85 (0.53,0.99) | 4 | 4.4 |
| Wantai Total Ig S/RBD | Total Ig | Spike | 0.56 (0.35,0.78) | 15 | 6 |
| Roche Elecsys Total Ig S* | Total Ig | Spike | 0.99 (0.98,1.00) | 6 | 4.9 |
| Ortho VITROS Total Ig S | Total Ig | Spike | 0.98 (0.95,0.99) | 15 | 6 |
| Roche Elecsys Total Ig N | Total Ig | Nucleocapsid | 0.99 (0.99,1.00) | 15 | 6 |
| Siemens ADVIA IgG S1/RBD | IgG | Spike | 0.89 (0.79,0.95) | 15 | 5.9 |
| Beckman Coulter Access IgG S1/RBD | IgG | Spike | 0.95 (0.90,0.98) | 15 | 5.9 |
| EUROIMMUN IgG S1 Qual | IgG | Spike | 0.90 (0.82,0.96) | 15 | 6 |
| EUROIMMUN IgG S1 Quant* | IgG | Spike | 0.91 (0.80,0.97) | 11 | 4.8 |
| Abbott IgG S1 (ARCHITECT) | IgG | Spike | 0.83 (0.70,0.93) | 15 | 6 |
| Abbott IgG S1 (Alinity) | IgG | Spike | 0.83 (0.70,0.93) | 15 | 6 |
| DiaSorin Trimeric IgG S | IgG | Spike | 0.90 (0.82,0.96) | 15 | 5.9 |
| Bio-Rad BioPlex MPX IgG RBD** | IgG | Spike | 0.95 (0.91,0.98) | 15 | 6 |
| Bio-Rad BioPlex MPX IgG S1** | IgG | Spike | 0.94 (0.88,0.98) | 15 | 6 |
| Bio-Rad BioPlex MPX IgG S2** | IgG | Spike | 0.94 (0.88,0.97) | 15 | 6 |
| Bio-Rad BioPlex MPX IgG N** | IgG | Nucleocapsid | 0.90 (0.82,0.96) | 15 | 6 |
| EUROIMMUN IgG N | IgG | Nucleocapsid | 0.81 (0.66,0.92) | 15 | 6 |
| Abbott IgG N (ARCHITECT) | IgG | Nucleocapsid | 0.91 (0.82, 0.96) | 15 | 6 |
| Abbott IgG N (Alinity) | IgG | Nucleocapsid | 0.86 (0.74,0.94) | 15 | 6 |
| Diazyme DZ-Lite IgG N/S | IgG | Multiantigen | 0.96 (0.91,0.98) | 15 | 6 |
| EUROIMMUN IgA S1 | IgA | Spike | 0.92 (0.85,0.97) | 14 | 5.9 |

**Appendix Table 2:** Intraclass correlation coefficients based on blinded replicate sample testing, reflecting the proportion of total variance that is between-sample rather than within-sample variability. S, spike protein; RBD, receptor binding domain; N, nucleocapsid; Ag, antigen; Ab, antibody; Ig, immunoglobulin. See Table 1 for assay details. N is the number of unique specimens (groups) included in the analysis, k is the number of measurements per group. These numbers are smaller than 15 and smaller than 6, respectively, in cases where some number of measurements fell outside the measurement range of the assay.

* Results falling outside the primary measurement range excluded.

** On-board dilutions were used to estimate reactivity in specimens where initial results fell outside the primary measurement range.

| **Blinded replicate sample** | **Sample 1** | **Sample 2** | **Sample 3** | **Sample 4** | **Sample 5** | **Sample 6** | **Sample 7** | **Sample 8** | **Sample 9** | **Sample 10** | **Sample 11** | **Sample 12** | **Sample 13** | **Sample 14** | **Sample 15** |
| --- | --- | --- | --- | --- | --- | --- | --- | --- | --- | --- | --- | --- | --- | --- | --- |
| **EUROIMMUN IgG S1 Qual** | 29.8 | 6 | 90.2 | 7.2 | 6.9 | 2.1 | 15.8 | 14.3 | 9.1 | 6.7 | 8.1 | 8.5 | 7.6 | 7.7 | 9.4 |
| **EUROIMMUN IgG S1 Quant** | 39.7 | 8.8 | 140.7 | 15.5 | 12.4 | 6.3 | 24.3 | 26.5 | 20.9 | 7.9 | 0.5 | 0 | 0 | 0 | 0 |
| **EUROIMMUN IgG N SemiQuant** | 33.1 | 12.4 | 24.8 | 13.6 | 13.6 | 17.1 | 17.5 | 26.2 | 33.6 | 15.2 | 6.2 | 23.4 | 15.8 | 14.6 | 22.6 |
| **EUROIMMUN IgA S1 SemiQuant** | 19.3 | 23.6 | 53.6 | 12.6 | 9.1 | 6.5 | 26.9 | 18 | 51.8 | 8.4 | 8.5 | 16.5 | 8.8 | * | 23.1 |
| **Roche Elecsys Total Ig N Qual** | 84 | 4.7 | 2.8 | 4.1 | 1.9 | 1.7 | 4.4 | 2.6 | 4.8 | 10.8 | 3.5 | 7.4 | 7 | 3.5 | 4.3 |
| **Roche Elecsys Total Ig S Quant** | 60.6 | 4.4 | 14.4 | 4.7 | 7.9 | 0 | 5.6 | 0 | 0 | 0 | 0 | 0 | 0 | 0 | 0 |
| **DiaSorin Trimeric IgG S1/S2 Quant** | 21.1 | 37.9 | 18.3 | 15.4 | 21.6 | 18.9 | 37.2 | 33.6 | 15.7 | 22.5 | 11.1 | 20.9 | 42.2 | 11 | 24.2 |
| **Siemens ADVIA Total Ig S1/RBD Qual** | 12.3 | 25.8 | 21.4 | 0 | 0 | 0 | 1.9 | 0 | 0 | 0 | 0 | 0 | 0 | 0 | 0 |
| **Siemens ADVIA IgG S1/RBD Quant** | 24.8 | 39.8 | 20.8 | 17.1 | 21.5 | 20.6 | 39.7 | 35.4 | 8.9 | 17.4 | 19.4 | 23 | 47.9 | 7.3 | 29.9 |
| **Abbott IgG N Qual (Alinity)** | 70.7 | 17.7 | 6.6 | 9.1 | 6.4 | 8.9 | 19.7 | 31.1 | 35.8 | 7.5 | 8.9 | 13.1 | 9.4 | 12.1 | 13 |
| **Abbott IgG S1 Quant (Alinity)** | 62.5 | 19.1 | 28.4 | 19 | 13 | 19.7 | 40.1 | 54.7 | 15.4 | 20.4 | 22.1 | 37.2 | 16 | 14.6 | 51.1 |
| **Abbott IgG N Qual (ARCHITECT)** | 66.5 | 15.5 | 6.1 | 11.6 | 9.5 | 5.4 | 18.9 | 33.3 | 7.7 | 8.3 | 2 | 14.6 | 4.4 | 3.5 | 2.3 |
| **Abbott IgG S1 Quant (ARCHITECT)** | 55.4 | 21.8 | 22.6 | 15.7 | 20.2 | 12.2 | 43 | 53.4 | 12.7 | 19.4 | 22.4 | 28.1 | 14.2 | 16.4 | 50.6 |
| **Bio-Rad BioPlex MPX IgG RBD** | 21.4 | 29.3 | 19 | 7.2 | 11.8 | 6.5 | 18.6 | 16.7 | 15.9 | 15.9 | 5.6 | 17.8 | 22.2 | 10.4 | 18.4 |
| **Bio-Rad BioPlex MPX IgG S1** | 24 | 34.9 | 20.9 | 6.3 | 10.4 | 7.3 | 18.4 | 20.7 | 15.9 | 13.3 | 5.4 | 21.8 | 25.6 | 10.8 | 21 |
| **Bio-Rad BioPlex MPX IgG S2** | 21.1 | 22.3 | ** | 5 | 12.9 | 11.9 | 19 | 14.8 | 10.8 | 13.3 | 11.7 | 20.5 | 14.2 | 10 | 19 |
| **Bio-Rad BioPlex MPX IgG N*** | 25.7 | 30.9 | 22 | 5.1 | 12.7 | 8.6 | 20.1 | 21.3 | 8.1 | 13 | 6.2 | 20.1 | 16.1 | 15 | 23.8 |
| **Bio-Rad Platelia Total Ig N SemiQuant** | 24.4 | 11.3 | 6.6 | 0 | 0 | 0 | 0 | 0 | 0 | 0 | 0 | 0 | 0 | 0 | 0 |
| **Beckman Coulter Access IgG S1/RBD** | 9.5 | 23.3 | 11.3 | 19.2 | 25 | 17.2 | 16.9 | 44.7 | 12.9 | 12.6 | 16.7 | 17 | 21.5 | 9.8 | 8.4 |
| **Diazyme DZ-Lite IgG N/S** | 18.6 | 22.4 | 22.6 | 22.3 | 24.3 | 30.8 | 27.1 | 76.5 | 8.8 | 17 | 25.2 | 33.3 | 14.4 | 37.7 | 14.1 |
| **Wantai Total Ig S/RBD** | 23.5 | 8.2 | 24.7 | 13.2 | 13 | 11.7 | 11.8 | 10.8 | 14.2 | 77.6 | 39.3 | 9.3 | 4.8 | 7.6 | 9.5 |
| **Ortho VITROS Total Ig S** | 11 | 11.3 | 7.3 | 7.3 | 4.4 | 7.2 | 14.6 | 3.3 | 13.4 | 6.4 | 16.1 | 6.4 | 4.6 | 7.4 | 7.1 |
| *** Not tested** |  |  |  |  |  |  |  |  |  |  |  |  |  |  |  |
| **** Negative** |  |  |  |  |  |  |  |  |  |  |  |  |  |  |  |

**Appendix Table 3:** Coefficients of variation reflecting within-sample variability from testing of six blinded replicates of fifteen distinct specimens. S, spike protein; RBD, receptor binding domain; N, nucleocapsid; Ag, antigen; Ab, antibody; Ig, immunoglobulin. See Table 1 for assay details.

Assays with limited dynamic range show some specimens “maxing out” at the maximum of the measurement range, resulting in spuriously low coefficients of variation. These are inidicated in red.

* On-board dilutions used to extend the dynamic range of the assay.

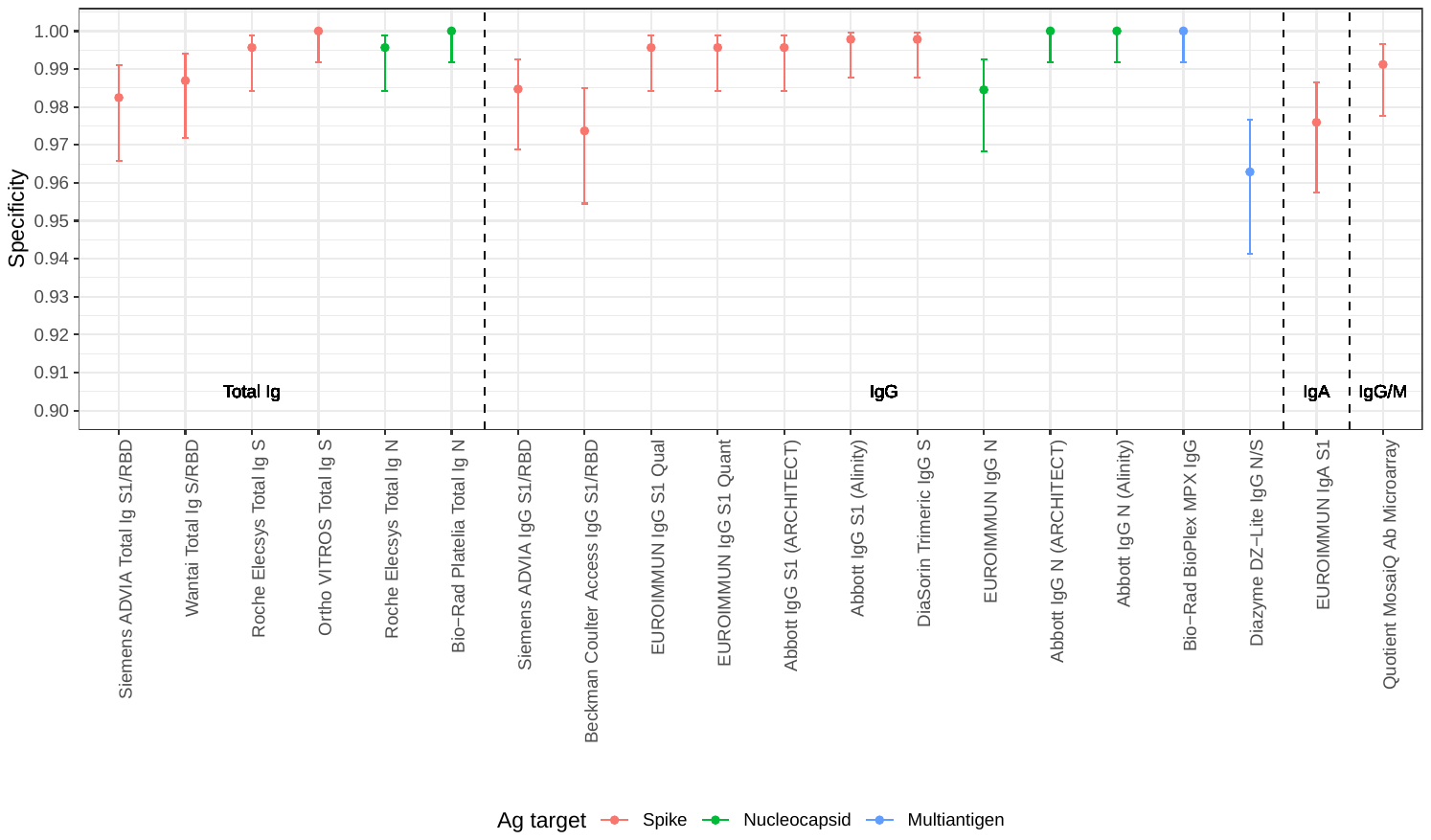

**Appendix Figure 1.** Specificity of of SARS-CoV-2 serological assays in pre-COVID-19 negative control specimens and early 2020 specimens. S, spike protein; RBD, receptor binding domain; N, nucleocapsid; Ag, antigen; Ab, antibody; Ig, immunoglobulin. See Table 1 for assay details.

Dots indicate point estimates and bars indicate Wilson score 95% confidence intervals.

**A**
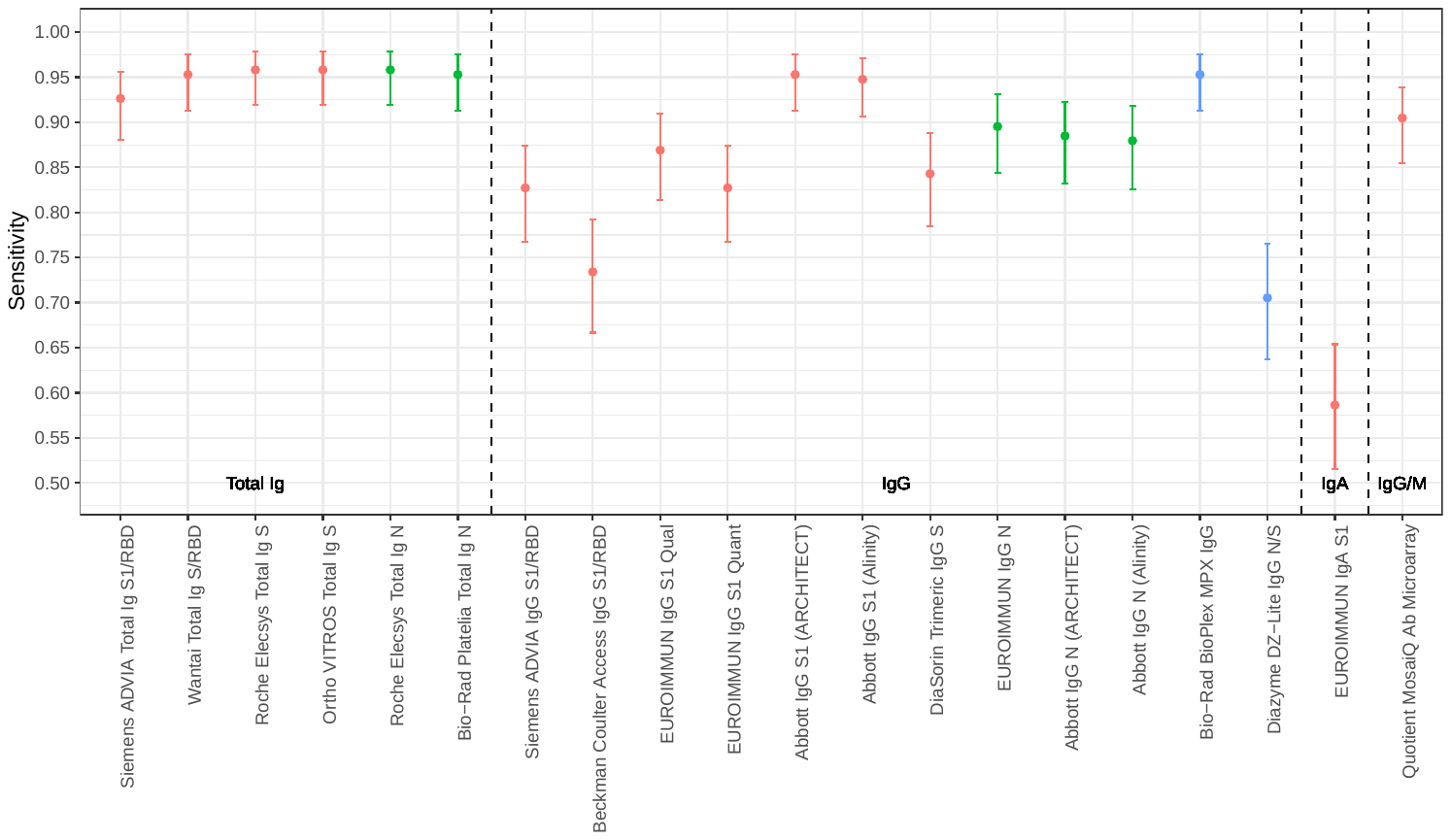

**B**
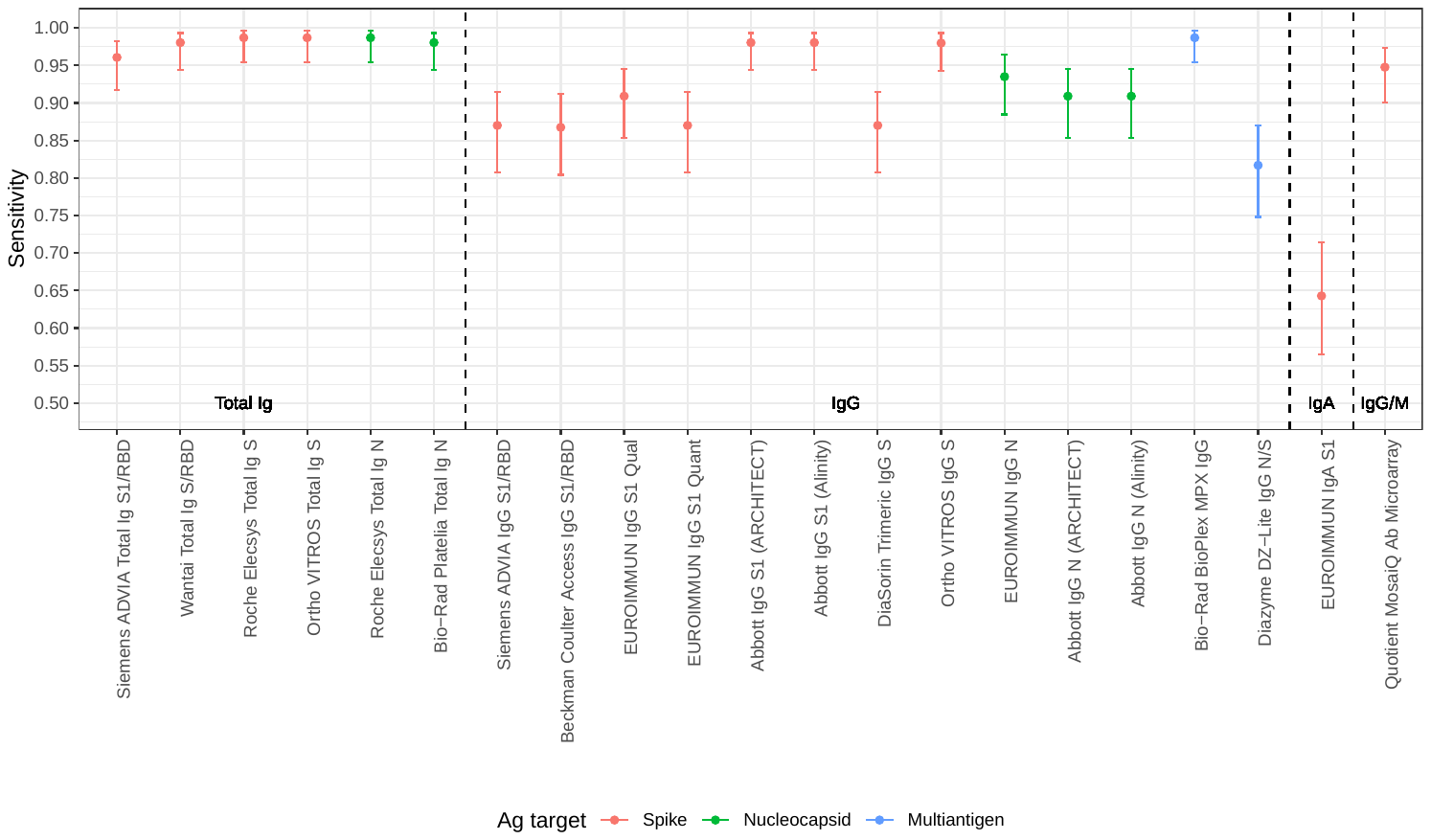

**C**
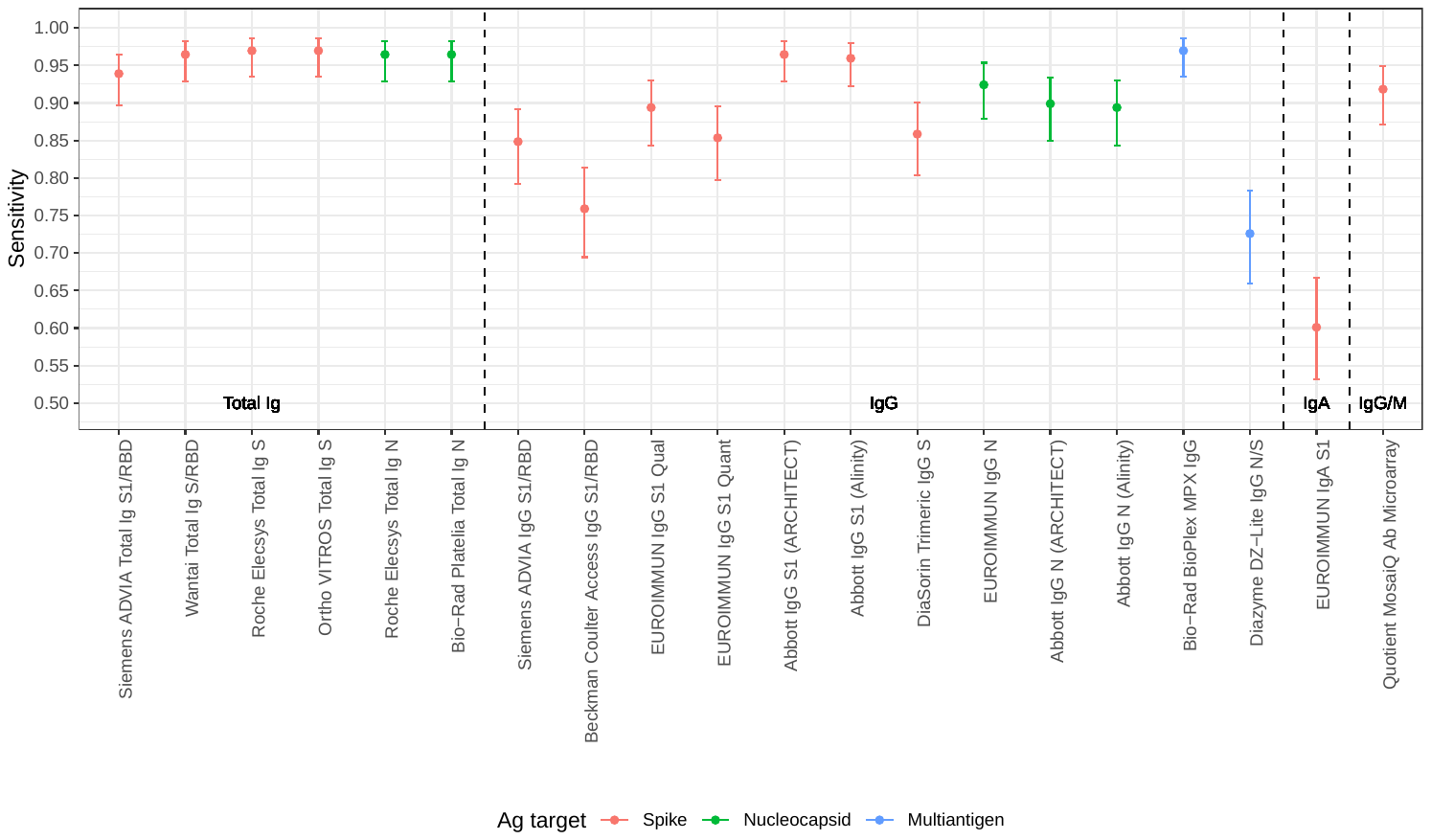

**Appendix Figure 2:** Sensitivity of SARS-CoV-2 serological assays using three reference definitions. *Secondary analysis with ‘equivocal’ results counted as negative.*

Panel A: Positivity defined by qualification as CCP donor (excluding purposely selected serosilents)

Panel B: Positivity defined by neutralizing activity measured by Broad PRNT

Panel C: Positivity defined by ‘operational standard’ (3 or more bAb assays reactive).

Dots indicate point estimates and bars indicate Wilson score 95% confidence intervals

*The Ortho VITROS IgG S assay is included only in panel B because it was tested on the subset of specimens with neutralization data available.* S, spike protein; RBD, receptor binding domain; N, nucleocapsid; Ag, antigen; Ab, antibody; Ig, immunoglobulin. See Table 1 for assay details.

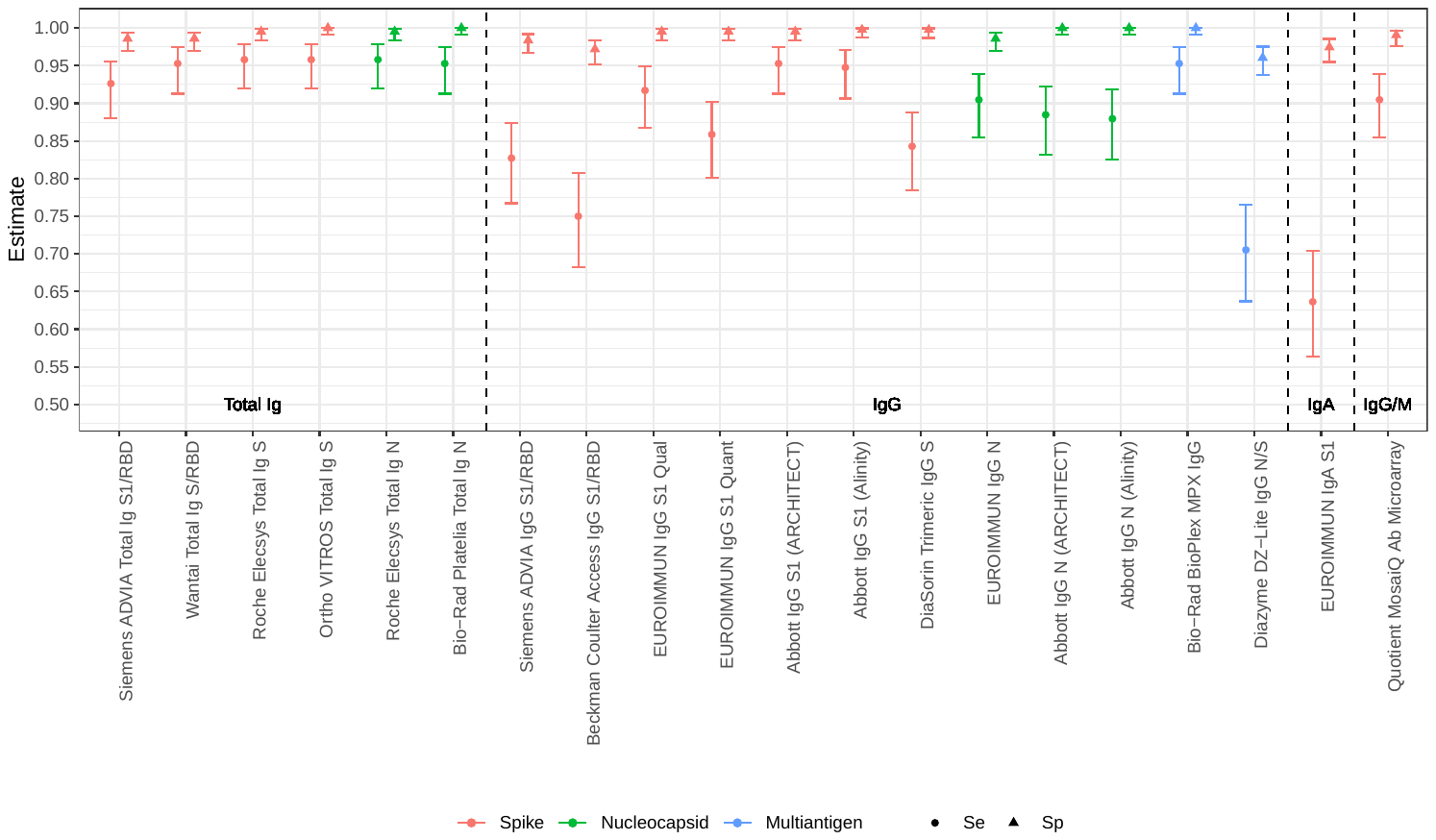

**Appendix Figure 3:** Sensitivity and specificity of SARS-CoV-2 serological assays with true positivity defined by reactivity on three or more bAb assays, and Specificity in 2019 pre-COVID-19 specimens. S, spike protein; RBD, receptor binding domain; N, nucleocapsid; Ag, antigen; Ab, antibody; Ig, immunoglobulin.

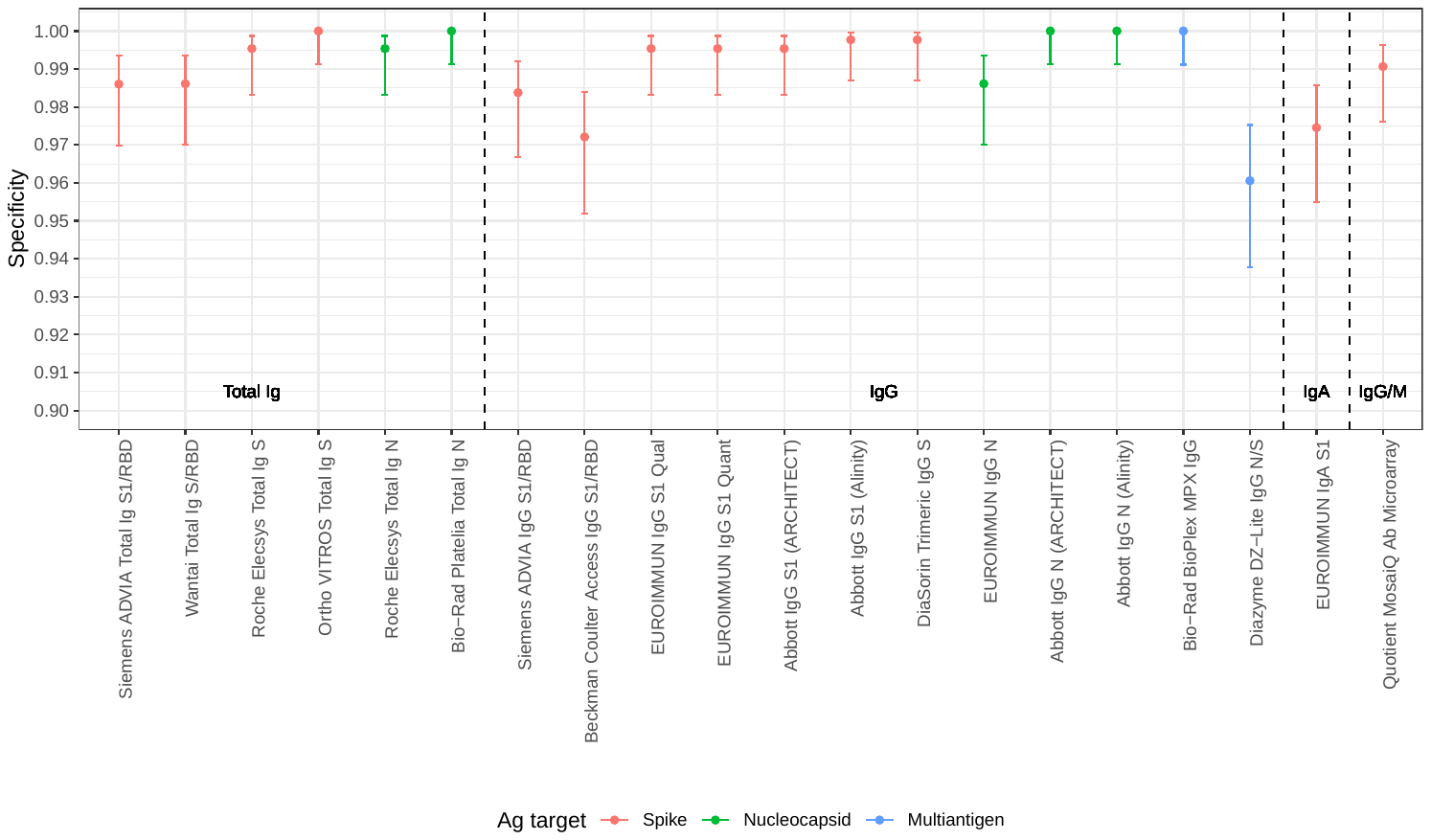

**Appendix Figure 4.** Specificity of SARS-CoV-2 serological assays in pre-COVID-19 negative control specimens. *Secondary analysis with ‘equivocal’ and ‘borderline’ results counted as negative.** S, spike protein; RBD, receptor binding domain; N, nucleocapsid; Ag, antigen; Ab, antibody; Ig, immunoglobulin.

* Some assays’ instructions for use define a range of signal intensities for which the interpretation is ‘equivocal’, with intensities above the upper limit of this range interpreted as ‘reactive’, and those below the lower limit as ‘non-reactive’.

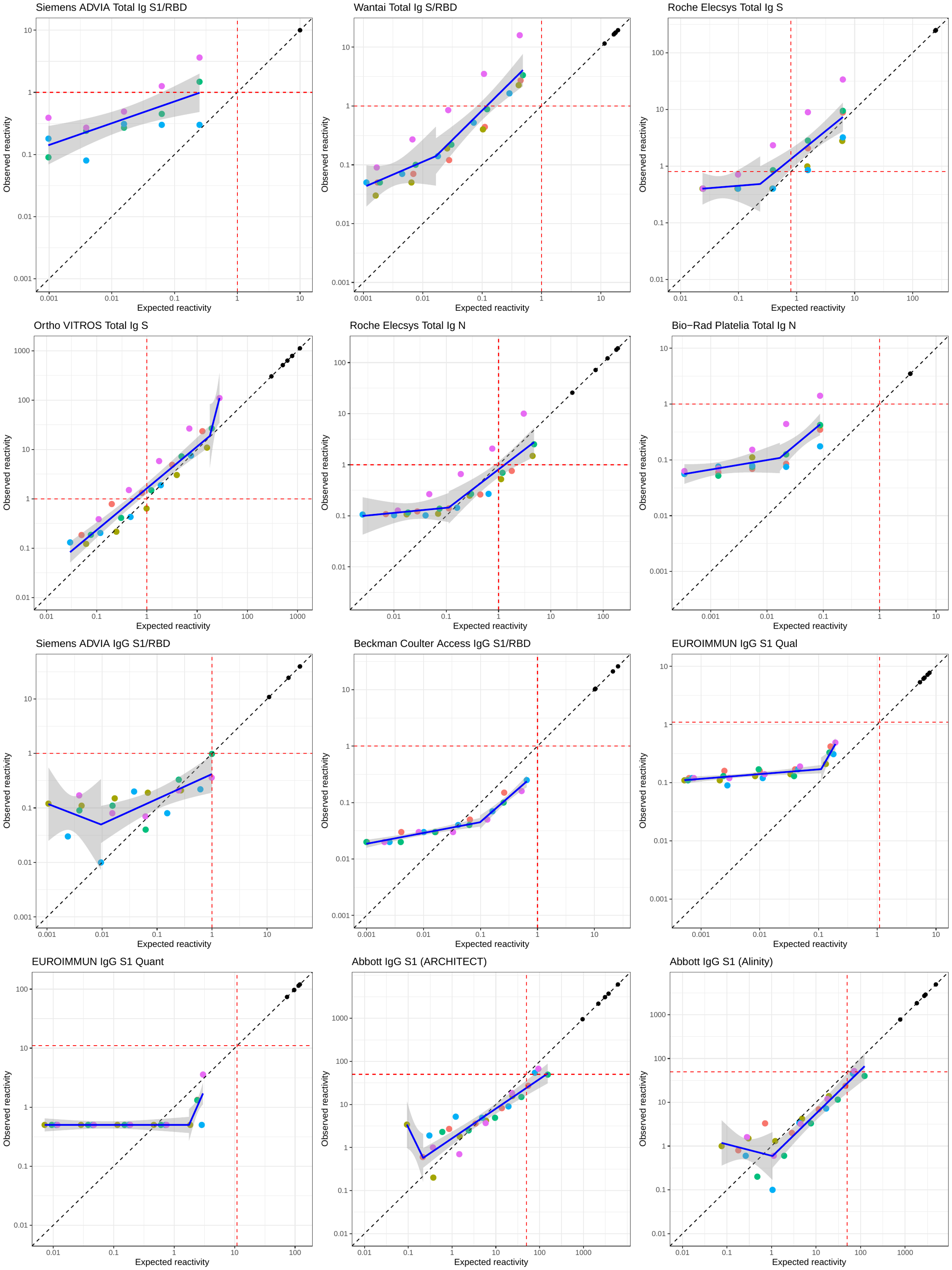

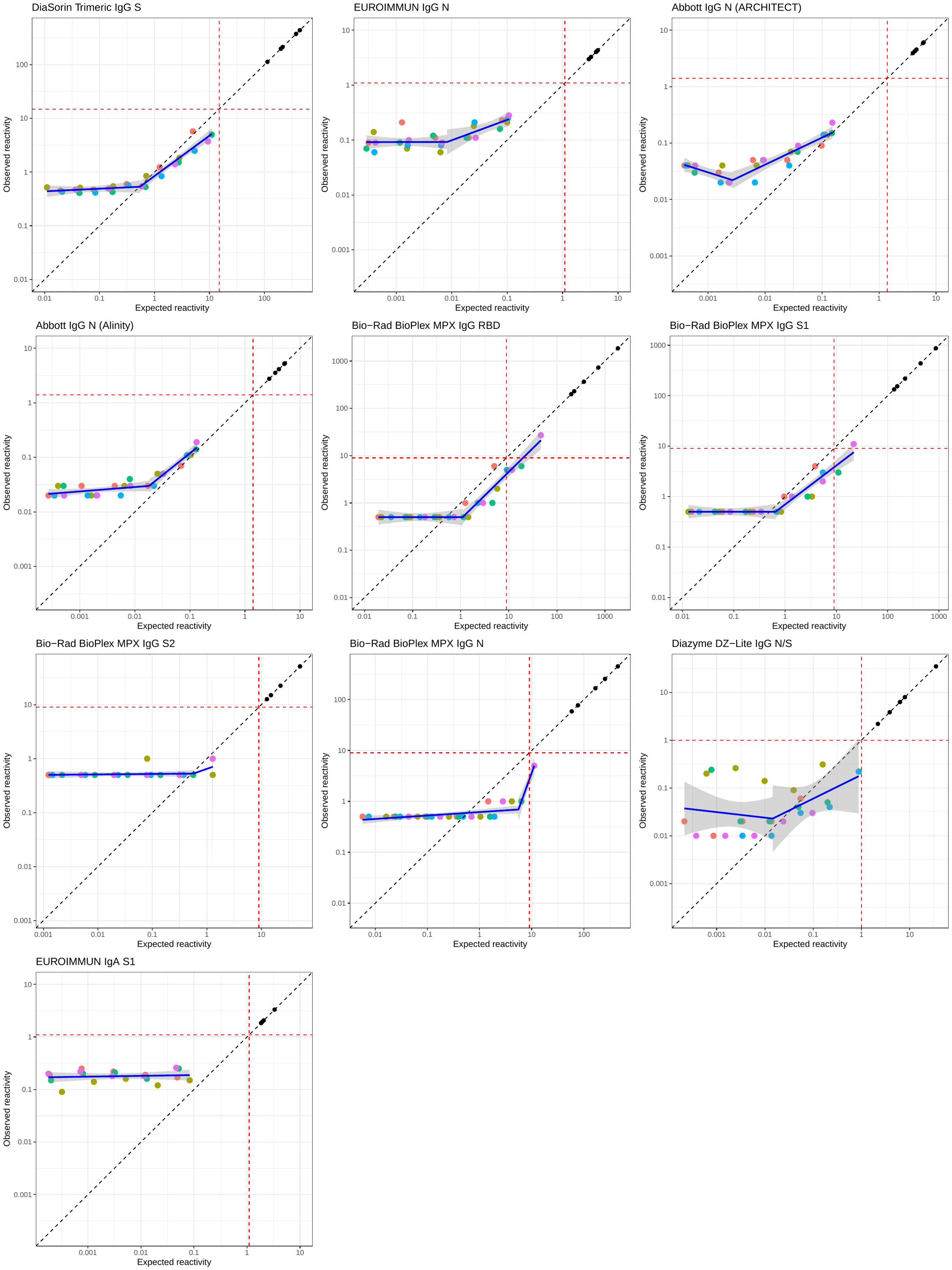

**Appendix Figure 5** Observed against expected signal intensity in five 4-fold serial dilutions of specimens with a range of neat Ab titers. S, spike protein; RBD, receptor binding domain; N, nucleocapsid; Ag, antigen; Ab, antibody; Ig, immunoglobulin. Expected reactivity is defined as the mean signal intensity measured over six replicates of the neat specimen divided by the dilution factor.

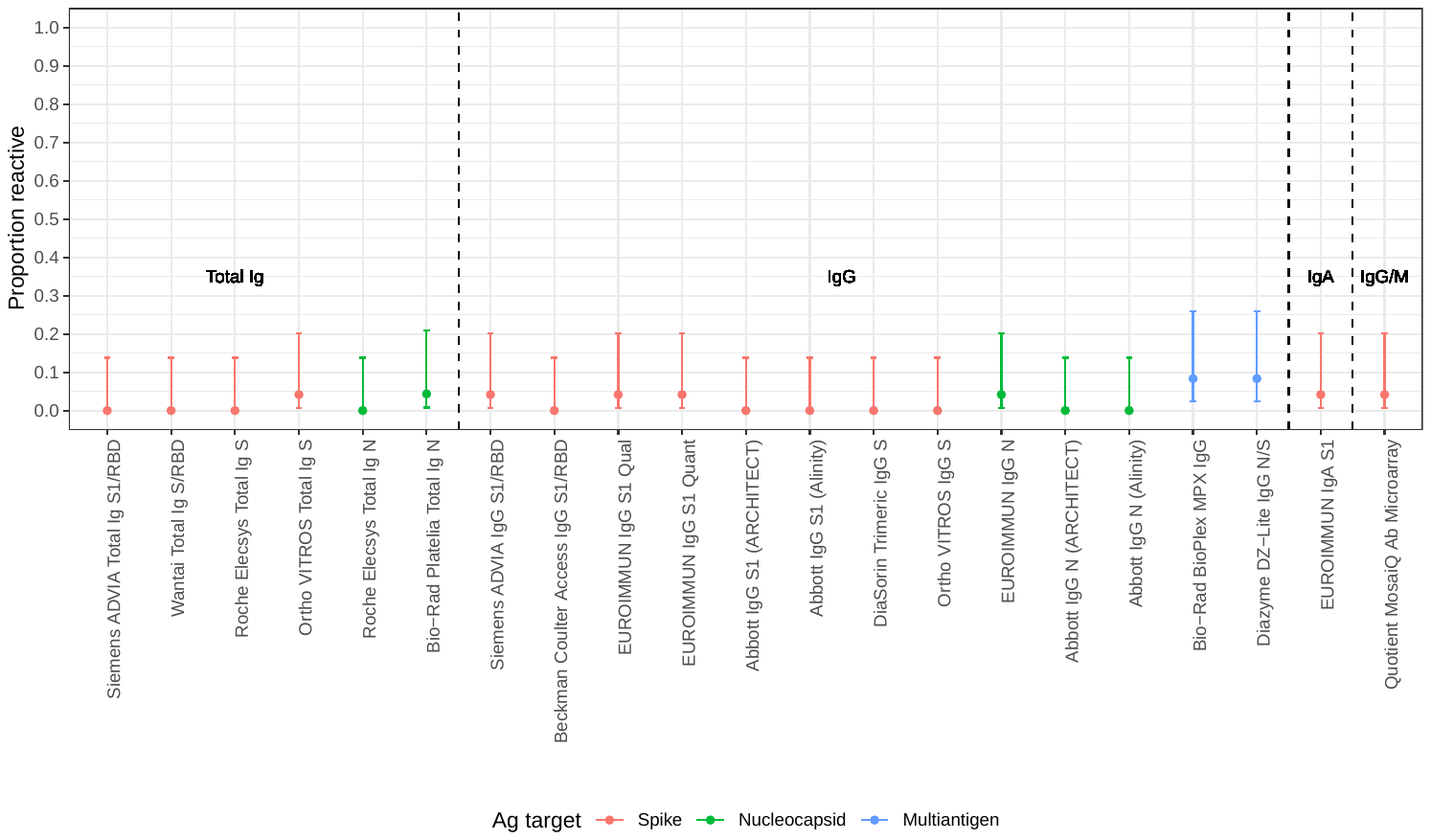

**Appendix Figure 6.** Proportion of “serosilent” CCP donors (non-reactive on Ortho VITROS Total Ig S during primary blood donor screening) detected by SARS-CoV-2 serological assays. S, spike protein; RBD, receptor binding domain; N, nucleocapsid; Ag, antigen; Ab, antibody; Ig, immunoglobulin.

**A**

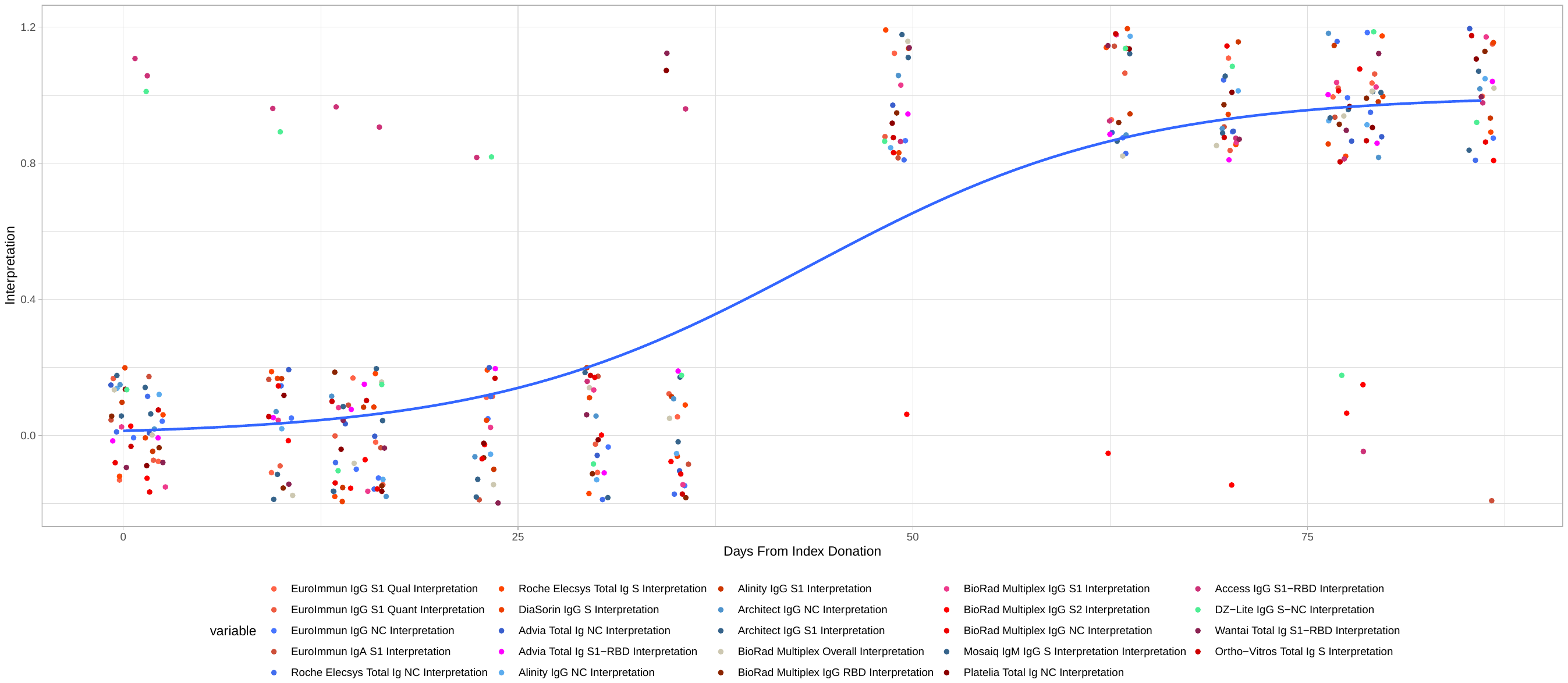

**B**
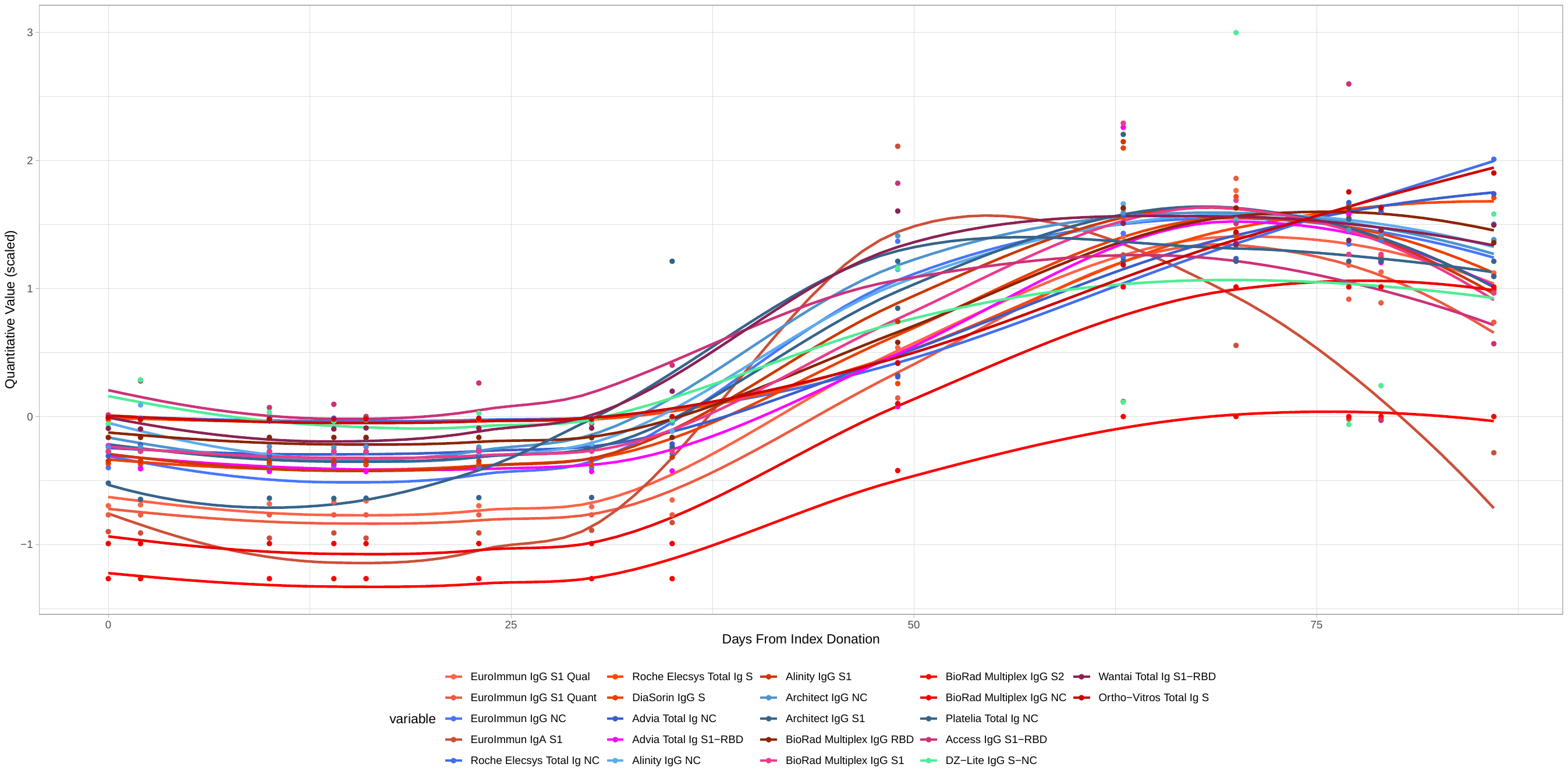

**Appendix Figure 7** Relative Timing of Seroconversion using a single seroconversion series without documented date of RT-PCR-based diagnosis. Legend explaining panel A and panel B

Panel A: Timing of reactivity by qualitative interpretation. The line indicates the probability of detection across assays based on regression analysis.

Panel B: Quantitative signal over time. Signal was rescaled so that 0 is each assay’s reactive/non-reactive cutoff and the range of signal intensities is on the arbitrary scale [-1,3]. S, spike protein; RBD, receptor binding domain; N, nucleocapsid; Ag, antigen; Ab, antibody; Ig, immunoglobulin.

**Appendix 1: Assessing trends in quantitative bAb signal intensity using longitudinal specimens from repeat COVID-19 Convalescent Plasma (CCP) donors**

Quantitative detection of binding antibodies was assessed by fitting linear mixed effects regression models with time since index donation the only predictor. The regression models included donor random effects (i.e. donor-specific slopes and intercepts) as well as fixed slopes and intercepts. Models were fit to log_10_-transformed and rescaled assay signal to render slope estimates comparable. Assay signal intensity was first log transformed and then rescaled to an arbitrary scale of 0 to 100, with zero the lowest value observed using a given assay, and 100 the highest value observed with that assay. We report the fixed (average) slopes with 95% confidence intervals in Figure 4A. For comparison, the slope in neutralizing Ab (nAb) titer, measured using the Broad institute plaque reduction neutralization test (PRNT) assay (and rescaled in the same way) is shown. For the single multiplexed assay included in the evaluation, each analyte is reported separately.

In assays with negative slopes that could be clearly distinguished from zero, i.e. where statistical evidence exists of waning reactivity, we estimated signal half-lives by fitting mixed effects regression models for log_2_-transformed assay signal, and used the fixed effect slopes to estimate half-lives using the following formula:

$$t_{1/2}=-\frac{1}{\beta}$$

with $t_{1/2}$ the half-life and $\beta$ the regression coefficient on days since index donation. We estimated half-lives only for assays where both point estimates and the upper bound of the 95% confidence interval on the slope were negative.
